# Benchmarking saliency methods for chest X-ray interpretation

**DOI:** 10.1101/2021.02.28.21252634

**Authors:** Adriel Saporta, Xiaotong Gui, Ashwin Agrawal, Anuj Pareek, Steven QH Truong, Chanh DT Nguyen, Van-Doan Ngo, Jayne Seekins, Francis G. Blankenberg, Andrew Y. Ng, Matthew P. Lungren, Pranav Rajpurkar

## Abstract

Saliency methods, which “explain” deep neural networks by producing heat maps that highlight the areas of the medical image that influence model prediction, are often presented to clinicians as an aid in diagnostic decision-making. Although many saliency methods have been proposed for medical imaging interpretation, rigorous investigation of the accuracy and reliability of these strategies is necessary before they are integrated into the clinical setting. In this work, we quantitatively evaluate seven saliency methods—including Grad-CAM, Grad-CAM++, and Integrated Gradients—across multiple neural network architectures using two evaluation metrics. We establish the first human benchmark for chest X-ray segmentation in a multilabel classification set up, and examine under what clinical conditions saliency maps might be more prone to failure in localizing important pathologies compared to a human expert benchmark. We find that (i) while Grad-CAM generally localized pathologies better than the other evaluated saliency methods, all seven performed significantly worse compared with the human benchmark; (ii) the gap in localization performance between Grad-CAM and the human benchmark was largest for pathologies that were smaller in size and had shapes that were more complex; (iii) model confidence was positively correlated with Grad-CAM localization performance. While it is difficult to know whether poor localization performance is attributable to the model or to the saliency method, our work demonstrates that several important limitations of saliency methods must be addressed before we can rely on them for deep learning explainability in medical imaging.

## Introduction

Deep learning has enabled automated medical imaging interpretation at the level of practicing experts in some settings^1–3^. While the potential benefits of automated diagnostic models are numerous, lack of model interpretability in the use of “black-box” deep neural networks (DNNs) represents a major barrier to clinical trust and adoption^4–6^. In fact, it has been argued that the European Union’s recently adopted General Data Protection Regulation (GDPR) affirms an individual’s right to an explanation in the context of automated decision-making^7^. Although the importance of DNN interpretability is widely acknowledged and many techniques have been proposed, little emphasis has been placed on how best to quantitatively evaluate these explainability methods^8^.

One type of DNN interpretation strategy widely used in the context of medical imaging is based on saliency (or pixel-attribution) methods^9–12^. Saliency methods produce heat maps highlighting the areas of the medical image that most influenced the DNN’s prediction. Since saliency methods provide post-hoc interpretability of models that are never exposed to bounding box annotations or pixel-level segmentations during training, they are particularly useful in the context of medical imaging where ground-truth segmentations can be especially time-consuming and expensive to obtain. The heat maps help to visualize whether a DNN is concentrating on the same regions of a medical image that a human expert would focus on, rather than concentrating on a clinically irrelevant part of the medical image or even on confounders in the image^13–15^. Saliency methods have been widely used for a variety of medical imaging tasks and modalities including, but not limited to, visualizing the performance of a convolutional neural network (CNN) in predicting (1) myocardial infarction^16^ and hypoglycemia^17^ from electrocardiograms, (2) visual impairment^18^, refractive error^19^, and anaemia^20^ from retinal photographs, (3) long-term mortality^21^ and tuberculosis^22^ from chest X-ray (CXR) images, and (4) appendicitis^23^ and pulmonary embolism^24^ on computed tomography scans. However, recent work has shown that saliency methods used to validate model predictions can be misleading in some cases and may lead to increased bias and loss of user trust in high-stakes contexts such as healthcare^25–28^. Therefore, a rigorous investigation of the accuracy and reliability of these strategies is necessary before they are integrated into the clinical setting^29^.

In this work, we perform a systematic evaluation of seven common saliency methods in medical imaging (Grad-CAM^30^, Grad-CAM++^31^, Integrated Gradients^32^, Eigen-CAM^41^, DeepLIFT^42^, Layer-Wise Relevance Propagation^43^, and Occlusion^44^) using three common CNN architectures (DenseNet121^33^, ResNet152^34^, Inception-v4^35^). In doing so, we establish the first human benchmark in CXR segmentation by collecting radiologist segmentations for 10 pathologies using CheXpert, a large publicly available CXR dataset^36^. To compare saliency method segmentations with expert segmentations, we use two metrics to capture localization accuracy: (1) *mean Intersection over Union*, a metric that measures the overlap between the saliency method segmentation and the expert segmentation, and (2) *hit rate*, a less strict metric than mIoU that does not require the saliency method to locate the full extent of a pathology. We find that (1) while Grad-CAM generally localizes pathologies more accurately than the other evaluated saliency methods, all seven perform significantly worse compared with a human radiologist benchmark (although it is difficult to know whether poor localization performance is attributable to the model or to the saliency method); (2) the gap in localization performance between Grad-CAM and the human benchmark is largest for pathologies that are smaller in size and have shapes that are more complex; (3) model confidence is positively correlated with Grad-CAM localization performance. We publicly release a development dataset of expert segmentations, which we call CheXlocalize, to facilitate further research in DNN explainability for medical imaging.

## Results

### Framework for evaluating saliency methods

Seven methods were evaluated—Grad-CAM, Grad-CAM++, Integrated Gradients, Eigen-CAM, DeepLIFT, Layer-Wise Relevance Propagation (LRP), and Occlusion—in a multi-label classification setup on the CheXpert dataset (Fig. 1a). We ran experiments using three CNN architectures previously used on CheXpert: DenseNet121, ResNet152, and Inception-v4. For each combination of saliency method and model architecture, we trained and evaluated an ensemble of 30 CNNs (see Methods for ensembling details). We then passed each of the CXRs in the dataset’s holdout test set into the trained ensemble model to obtain image-level predictions for the following 10 pathologies: Airspace Opacity, Atelectasis, Cardiomegaly, Consolidation, Edema, Enlarged Cardiomediastinum, Lung Lesion, Pleural Effusion, Pneumothorax, and Support Devices. Of the 14 observations labeled in the CheXpert dataset, Fracture and Pleural Other were not included in our analysis because they had low prevalence in our test set (fewer than 10 examples); Pneumonia was not included because it is a clinical (as opposed to a radiological) diagnosis; and No Finding was not included because it is not applicable to evaluating localization performance. For each CXR, we used the saliency method to generate heat maps, one for each of the 10 pathologies, and then applied a threshold to each heat map to produce binary segmentations (top row, Fig. 1a). Thresholding is determined per pathology using Otsu’s method^37^, which iteratively searches for a threshold value that maximizes inter-class pixel intensity variance. We also conducted a second thresholding scheme in which we iteratively search for a threshold value that maximizes per pathology mIoU on the validation set. There are no statistically significant differences between the two thresholding schemes when compared against the human benchmark (see Extended Data Fig. 1). Additionally, to calculate the hit rate evaluation metric (described below), we extracted the pixel in the saliency method heat map with the largest value as the single most representative point on the CXR for that pathology.

**Fig. 1.**
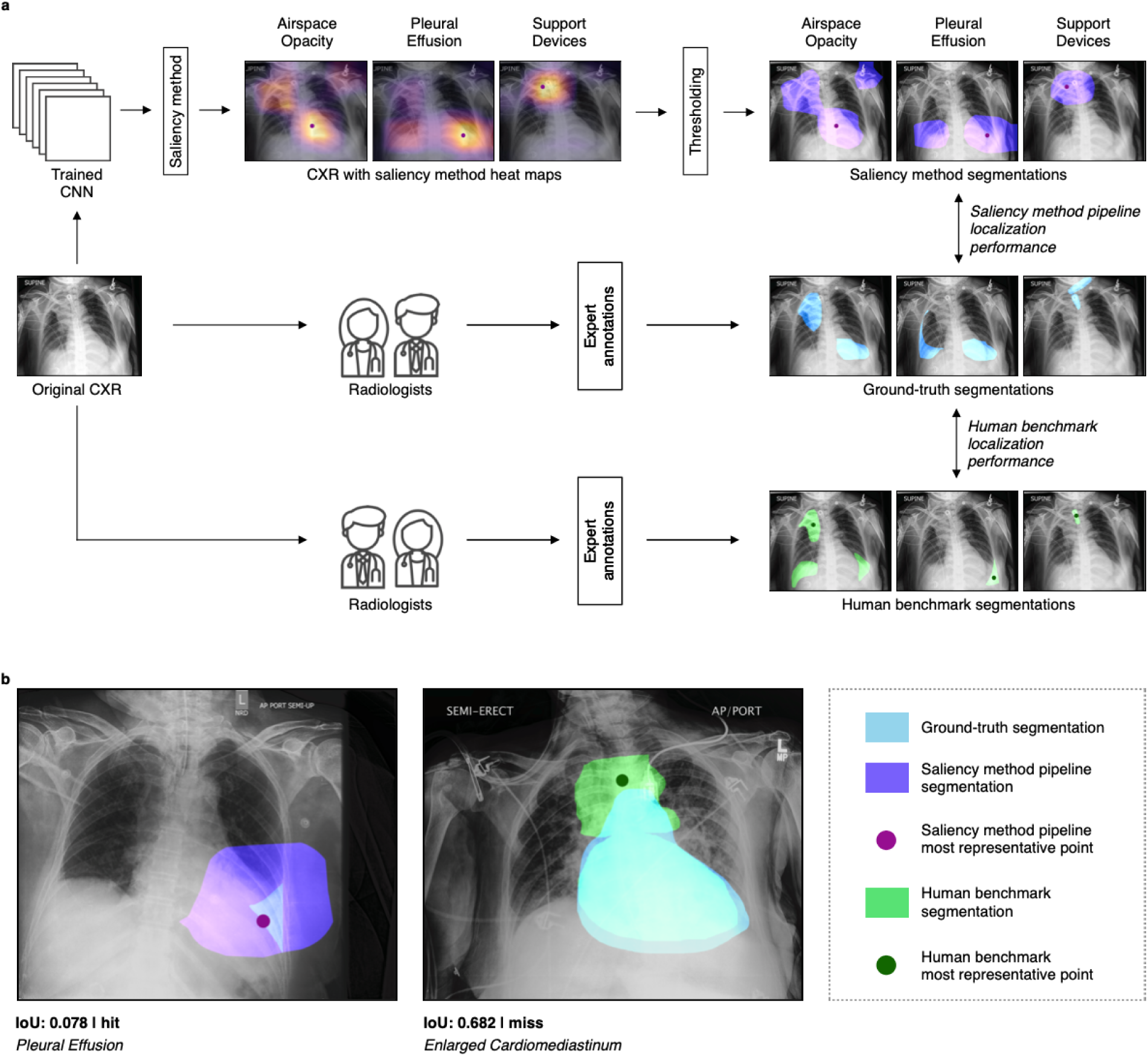
Framework for evaluating saliency methods. **a**, Top row left: a CXR image from the holdout test set is passed into an ensemble CNN trained only on CXR images and their corresponding pathology task labels. Saliency method is used to generate 10 heat maps for the example CXR, one for each task. The pixel in the heat map with the largest value is determined to be the single most representative point on the CXR for that pathology. There are three pathologies present in this CXR (Airspace Opacity, Pleural Effusion, and Support Devices). Top row right: a threshold is applied to the heat maps to produce binary segmentations for each present pathology. Middle row: Two board-certified radiologists were asked to segment the pathologies that were present in the CXR as determined by the dataset’s ground-truth labels. Saliency method pipeline annotations are compared to these ground-truth annotations to determine “saliency method pipeline localization performance”. Bottom row: Three board-certified radiologists (separate from those in middle row) were also asked to segment the pathologies that were present in the CXR as determined by the dataset’s ground-truth labels. In addition, these radiologists were asked to locate the single point on the CXR that was most representative of each present pathology. These benchmark annotations are compared to the ground-truth annotations to determine “human benchmark localization performance”. **b**, Left: CXR with ground-truth and saliency method annotations for Pleural Effusion. The segmentations have a low overlap (IoU is 0.078), but pointing game is a “hit” since the saliency method’s most representative point is inside of the ground-truth segmentation. Right, CXR with ground-truth and human benchmark annotations for Enlarged Cardiomediastinum. The segmentations have a high overlap (IoU is 0.682), but pointing game is a “miss” since saliency method’s most representative point is outside of the ground-truth segmentation.

We obtained two independent sets of pixel-level CXR segmentations on the holdout test set: ground-truth segmentations drawn by two board-certified radiologists (middle row, Fig. 1a) and human benchmark segmentations drawn by a separate group of three board-certified radiologists (bottom row, Fig. 1a). The human benchmark segmentations and the saliency method segmentations were compared with the ground-truth segmentations to establish the human benchmark localization performance and the saliency method localization performance, respectively. Additionally, for the hit rate evaluation metric, the radiologists who drew the benchmark segmentations were also asked to locate a single point on the CXR that was most representative of the pathology at hand (see Supplementary Figs. S1 through S11 for detailed instructions given to the radiologists). Note that the human benchmark localization performance demonstrates interrater variability, and we use it as a reference when evaluating saliency method pipelines.

We used two evaluation metrics to compare segmentations (Fig. 1b). Our primary metric, *mean Intersection over Union* (mIoU), measures how much, on average, either the saliency method or benchmark segmentations overlapped with the ground-truth segmentations. Our secondary metric, *hit rate*, is a less strict metric that does not require the saliency method or benchmark annotators to locate the full extent of a pathology. Hit rate is based on the pointing game setup^38^, in which credit is given if the most representative point identified by the saliency method or the benchmark annotators lies within the ground-truth segmentation. A “hit” indicates that the correct region of the CXR was located regardless of the exact bounds of the binary segmentations. Localization performance is then calculated as the hit rate across the dataset^39^. We report the mean of these metrics (mIoU and hit rate) over 1,000 bootstrap replicates on the test set, along with the 95% confidence intervals using the 2.5^th^ and 97.5^th^ percentiles of the empirical distribution^40^. In addition to mIoU, we report the test set precision, recall/sensitivity, and specificity values of the saliency method pipeline and the human benchmark segmentations to measure segmentation overlap (Extended Data Fig. 2).

### Evaluating localization performance

In order to compare the localization performance of the saliency methods with the human benchmark, we first used Grad-CAM, Grad-CAM++, and Integrated Gradients to run eighteen experiments, one for each combination of saliency method (Grad-CAM, Grad-CAM++, or Integrated Gradients) and CNN architecture (DenseNet121, ResNet152, or Inception-v4) using one of the two evaluation metrics (mIoU or hit rate) (see Extended Data Fig. 3). We also ran experiments to evaluate the localization performances of DenseNet121 with Eigen-CAM, DeepLIFT, LRP, and Occlusion. We found that Grad-CAM with DenseNet121 generally demonstrated better localization performance across pathologies and evaluation metrics than the other combinations of saliency method and architecture. Accordingly, we compared Grad-CAM with DenseNet121 (“saliency method pipeline”) with the human benchmark using both mIoU and hit rate. See Table 1 for localization performance on the test set of all seven saliency methods using DenseNet121. The localization performance for each pathology is reported on the true positive slice of the dataset (for mIoU, the true positive slice contains CXRs with both saliency method/human benchmark segmentations and also ground-truth segmentations; for hit rate, the true positive slice contains CXRs with both the most representative point identified by the saliency method/human benchmark and also the ground-truth segmentations). Localization performance was calculated this way so that saliency methods were not penalized by DNN classification error: while the benchmark radiologists were provided with ground-truth labels when annotating the dataset, saliency method segmentations were created based on labels predicted by the model. (See Extended Data Fig. 4 for saliency method pipeline test set localization performance on the full dataset using mIoU.)

**Table 1.** Test set localization performance of saliency methods using DenseNet121.

|  |  |  |  |  |  |  |  |
| --- | --- | --- | --- | --- | --- | --- | --- |
| Pathology | Grad-CAM | Grad-CAM++ | Integrated Gradients | Eigen-CAM | DeepLIFT | LRP | Occlusion |
| mIoU |  |  |  |  |  |  |  |
| Airspace Opacity | 0.248 | 0.234 | 0.123 | 0.293 | 0.111 | 0.112 | 0.242 |
| Atelectasis | 0.254 | 0.245 | 0.116 | 0.267 | 0.126 | 0.109 | 0.250 |
| Cardiomegaly | 0.452 | 0.346 | 0.160 | 0.379 | 0.167 | 0.150 | 0.312 |
| Consolidation | 0.408 | 0.296 | 0.177 | 0.332 | 0.088 | 0.099 | 0.212 |
| Edema | 0.362 | 0.388 | 0.073 | 0.370 | 0.059 | 0.047 | 0.347 |
| Enlarged Cardiom. | 0.379 | 0.400 | 0.154 | 0.372 | 0.109 | 0.117 | 0.363 |
| Lung Lesion | 0.101 | 0.089 | 0.107 | 0.089 | 0.072 | 0.088 | 0.087 |
| Pleural Effusion | 0.235 | 0.195 | 0.088 | 0.249 | 0.090 | 0.082 | 0.215 |
| Pneumothorax | 0.213 | 0.216 | 0.077 | 0.218 | 0.084 | 0.066 | 0.214 |
| Support Devices | 0.163 | 0.133 | 0.099 | 0.116 | 0.086 | 0.052 | 0.126 |
| Hit rate |  |  |  |  |  |  |  |
| Airspace Opacity | 0.498 | 0.558 | 0.606 | 0.566 | 0.528 | 0.566 | 0.367 |
| Atelectasis | 0.501 | 0.621 | 0.520 | 0.530 | 0.415 | 0.468 | 0.343 |
| Cardiomegaly | 0.903 | 0.732 | 0.697 | 0.709 | 0.610 | 0.644 | 0.515 |
| Consolidation | 0.738 | 0.708 | 0.624 | 0.626 | 0.571 | 0.283 | 0.338 |
| Edema | 0.746 | 0.781 | 0.300 | 0.758 | 0.468 | 0.156 | 0.469 |
| Enlarged Cardiom. | 0.818 | 0.630 | 0.704 | 0.612 | 0.469 | 0.594 | 0.767 |
| Lung Lesion | 0.290 | 0.290 | 0.423 | 0.146 | 0.497 | 0.356 | 0.072 |
| Pleural Effusion | 0.507 | 0.347 | 0.332 | 0.439 | 0.408 | 0.283 | 0.291 |
| Pneumothorax | 0.392 | 0.489 | 0.801 | 0.195 | 0.801 | 0.697 | 0.297 |
| Support Devices | 0.355 | 0.364 | 0.491 | 0.216 | 0.598 | 0.264 | 0.189 |

We found that the saliency method pipeline demonstrated significantly worse localization performance on the test set when compared with the human benchmark using both mIoU (Fig. 2a) and hit rate (Fig. 2b) as an evaluation metric, regardless of model classification AUROC. For five of the 10 pathologies, the saliency method pipeline had a significantly lower mIoU than the human benchmark. For example, the saliency method pipeline had one of the highest AUROC scores of the 10 pathologies for Support Devices (0.969), but had among the worst localization performance for Support Devices when using both mIoU (0.163 [95% CI 0.154, 0.172]) and hit rate (0.355 [95% CI 0.303, 0.408]) as evaluation metrics. On two pathologies (Atelectasis and Consolidation) the saliency method pipeline significantly outperformed the human benchmark. On average, across all 10 pathologies, mIoU saliency method pipeline performance was 24.0% [95% CI 18.2%, 29.6%] worse than the human benchmark, with Lung Lesion displaying the largest gap in performance (76.2% [95% CI 59.1%, 87.5%] worse than the human benchmark) (Extended Data Fig. 5). Consolidation was the pathology on which the mIoU saliency method pipeline performance exceeded the human benchmark the most, by 128.1%. For seven of the 10 pathologies, the saliency method pipeline had a significantly lower hit rate than the human benchmark. On average, hit rate saliency method pipeline performance was 29.4% [95% CI 23.1%, 35.5%] worse than the human benchmark (Extended Data Fig. 6), with Lung Lesion again displaying the largest gap in performance (65.9% [95% CI 35.3%, 91.7%] worse than the human benchmark). The hit rate saliency method pipeline did not significantly outperform the human benchmark on any of the 10 pathologies; for the remaining three of the 10 pathologies, the hit rate performance differences between the saliency method pipeline and the human benchmark were not statistically significant. Therefore, while the saliency method pipeline significantly underperformed the human benchmark regardless of evaluation metric used, the average performance gap was larger when using hit rate as an evaluation metric than when using mIoU as an evaluation metric.

**Fig. 2.**
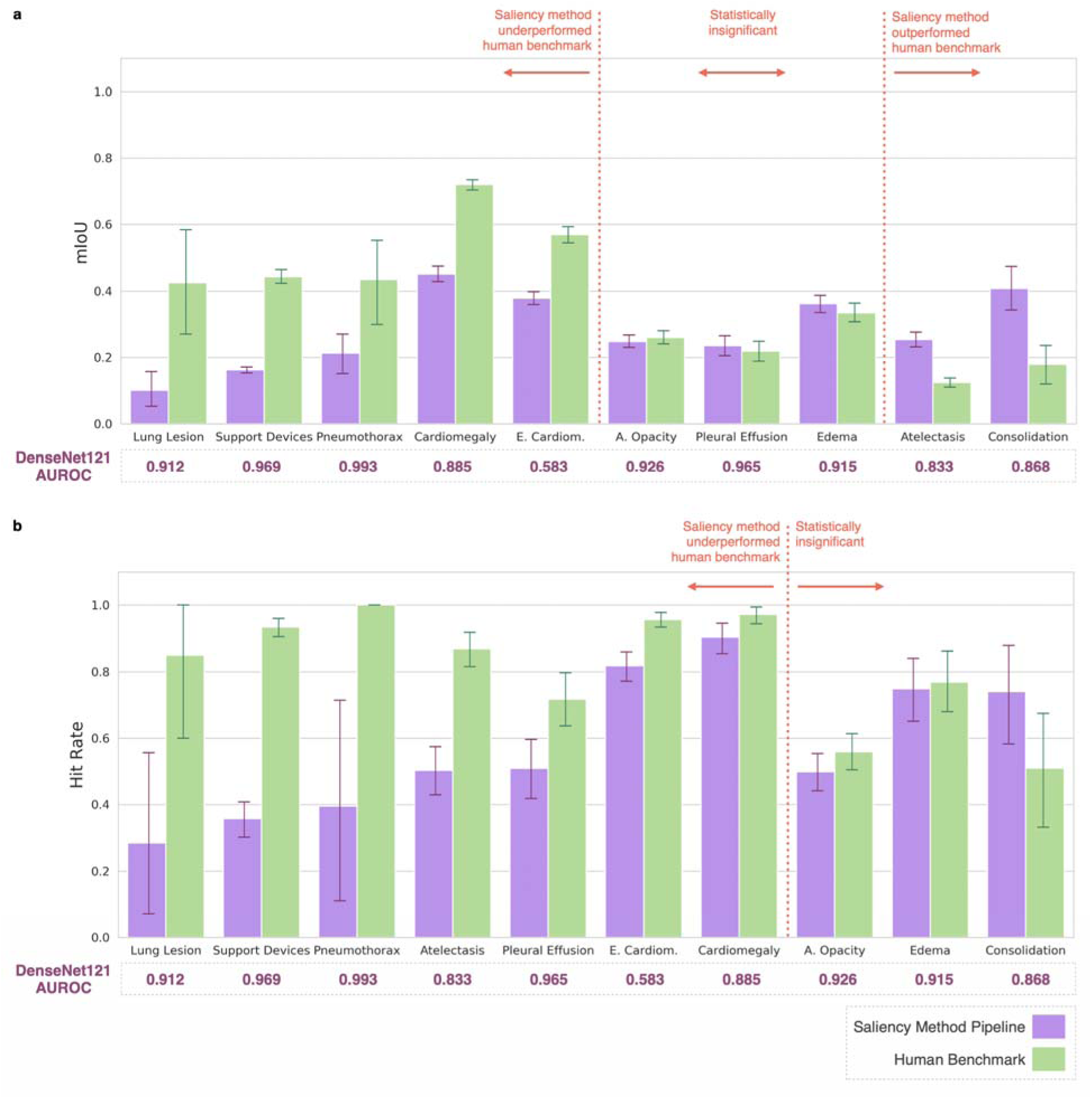
Evaluating localization performance. **a**, performances on the test set using mIoU. **b**, Comparing saliency method pipeline and human benchmark localization performances on the test set using hit rate. For both **a** and **b**, pathologies, along with their DenseNet121 AUROCs, are sorted on the x-axis first by statistical significance of percentage decrease from human benchmark mIoU/hit rate to saliency method pipeline mIoU/hit rate (high to low), and then by percentage decrease from human benchmark mIoU/hit rate to saliency method pipeline mIoU/hit rate (high to low).

We compared saliency method pipeline localization performance using an ensemble model to localization performance using the top performing single checkpoint for each pathology. We found that on the test set the single model has worse localization performance than the ensemble model for all pathologies when using mIoU and for six of the 10 pathologies when using hit rate (see Extended Data Fig. 7).

### Characterizing underperformance of saliency method pipeline

In order to better understand the underperformance of the saliency method pipeline localization, we first conducted a qualitative analysis with a radiologist by visually inspecting both the segmentations produced by the saliency method pipeline (Grad-CAM with DenseNet121) and the human benchmark segmentations. We found that, in general, saliency method segmentations fail to capture the geometric nuances of a given pathology, and instead produce coarse, low-resolution heat maps. Specifically, our qualitative analysis found that the performance of the saliency method was associated with three pathological characteristics (Fig. 3a): (1) *number of instances*: when a pathology had multiple instances on a CXR, the saliency method segmentation often highlighted one large confluent area, instead of highlighting each distinct instance of the pathology separately; (2) *size*: saliency method segmentations tended to be significantly larger than human expert segmentations, often failing to respect clear anatomical boundaries; (3) *shape complexity*: the saliency method segmentations for pathologies with complex shapes frequently included significant portions of the CXR where the pathology is not present.

**Fig. 3.**
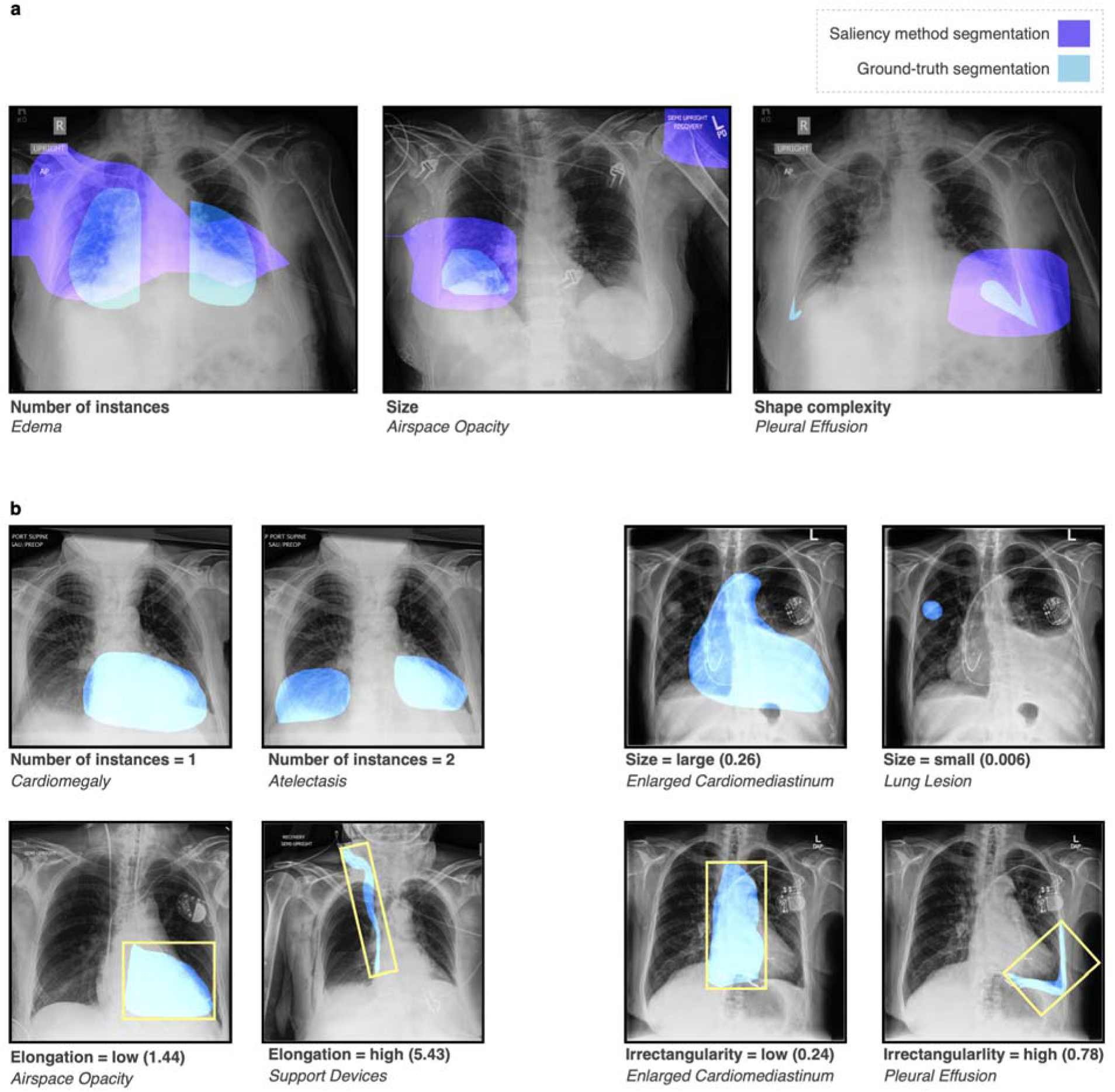
Characterizing underperformance of saliency method pipeline. **a**, Example CXRs that highlight the three pathological characteristics identified by our qualitative analysis: (1) Left, number of instances; (2) Middle, size; and (3) Right, shape complexity. **b**, Example CXRs with the four geometric features used in our quantitative analysis: (1) Top row left, number of instances; (2) Top row right, size = area of segmentation/area of CXR; (3) Bottom row left, elongation; and (4) Bottom row right, irrectangularity. Elongation and irrectangularity were calculated by fitting a rectangle of minimum area enclosing the binary mask (as indicated by the yellow rectangles). Elongation = maxAxis/minAxis. Irrectangularity = 1 - (area of segmentation/area of enclosing rectangle).

Informed by our qualitative analysis and previous work in histology^45^, we defined four geometric features for our quantitative analysis (Fig. 3b): (1) *number of instances* (for example, bilateral Pleural Effusion would have two instances, whereas there is only one instance for Cardiomegaly), (2) *size* (pathology area with respect to the area of the whole CXR), (3) *elongation* and (4) *irrectangularity* (the last two features measure the complexity of the pathology shape and were calculated by fitting a rectangle of minimum area enclosing the binary mask). See Extended Data Fig. 8 for the test set distribution of the four pathological characteristics across all 10 pathologies.

For each evaluation metric, we ran 8 simple linear regressions: four with the evaluation metric (IoU or hit/miss) of the saliency method pipeline (Grad-CAM with DenseNet121) as the dependent variable (to understand the relationship between the geometric features of a pathology and saliency method localization performance), and four with the difference between the evaluation metrics of the saliency method pipeline and the human benchmark as the dependent variable (to understand the relationship between the geometric features of a pathology and the gap in localization performance between the saliency method pipeline and the human benchmark). Each regression used one of the four geometric features as a single independent variable, and only the true positive slice was included in each regression. Each feature was normalized using min-max normalization and the regression coefficient can be interpreted as the effect of that geometric feature on the evaluation metric at hand. See Table 2 for coefficients from the regressions using both evaluation metrics on the test set, where we also report the 95% confidence interval and the Bonferroni corrected p-values based on Student’s t-distribution.

**Table 2.** Coefficients from regressions on geometric features of pathologies.

| Geometric feature<br>(independent variable) | Coefficient using<br>saliency method localization<br>(dependent variable) | Coefficient using localization difference<br>(human benchmark - saliency method)<br>(dependent variable) |
| --- | --- | --- |
| IoU |  |  |
| Number of instances | -0.115 (-0.220, -0.010) | -0.072 (-0.237, 0.094) |
| Size | 0.566 (0.526, 0.606) *** | -0.154 (-0.231, -0.076) *** |
| Elongation | -0.425 (-0.497, -0.354) *** | 0.476 (0.362, 0.589) *** |
| Irrectangularity | -0.256 (-0.292 -0.219) *** | 0.307 (0.249, 0.366) *** |
| Hit/Miss |  |  |
| Number of instances | -0.051 (-0.346, 0.244) | 0.470 (0.114, 0.825) * |
| Size | 1.269 (1.146, 1.391) *** | -0.944 (-1.104, -0.785) *** |
| Elongation | -0.849 (-1.053, -0.646) *** | 1.110 (0.865, 1.354) *** |
| Irrectangularity | -0.519 (-0.624, -0.415) *** | 0.689 (0.564, 0.815) *** |
| * p-value < 0.05, ** p-value < 0.01, *** p-value < 0.001 |  |  |

Our statistical analysis showed that as the size of a pathology increased, IoU saliency method localization performance improved (0.566 [95% CI 0.526, 0.606]). We also found that as elongation and irrectangularity increased, IoU saliency method localization performance worsened (elongation: −0.425 [95% CI −0.497, −0.354], irrectangularity: −0.256 [95% CI −0.292, −0.219]). We observed that the effects of these three geometric features were similar for hit/miss saliency method localization performance in terms of levels of statistical significance and direction of the effects. However, there was no evidence that the number of instances of a pathology had a significant effect on either IoU (−0.115 [95% CI −0.220, −0.010]) or hit/miss (−0.051 [95% CI −0.346, 0.244]) saliency method localization. Therefore, regardless of evaluation metric, saliency method localization performance suffered in the presence of pathologies that were small in size and complex in shape.

We found that these same three pathological characteristics—size, elongation, and irrectangularity—characterized the *gap* in IoU localization performance between saliency method and human benchmark. We observed that the *gap* in hit/miss localization performance was significantly characterized by all four geometric features (number of instances, size, elongation, and irrectangularity).

### Effect of model confidence on localization performance

We also conducted statistical analyses to determine whether there was any correlation between the model’s confidence in its prediction and saliency method pipeline test set localization performance (Table 3). We first ran a simple regression for each pathology using the model’s probability output as the single independent variable and using the saliency method IoU as the dependent variable. We then performed a simple regression that uses the same approach as above, but that includes all 10 pathologies. For each of the 11 regressions, we used the full dataset since the analysis of false positives and false negatives was also of interest. In addition to the linear regression coefficients, we also computed the Spearman correlation coefficients to capture any potential non-linear associations.

**Table 3.** IoU: Coefficients from regressions on model assurance.

| **Table 3 \| IoU: Coefficients from regressions on model assurance** |  |  |  |
| --- | --- | --- | --- |
| **Pathology** | **CXRs including all positives and false negatives (n)** | **Linear regression coefficient** | **Spearman correlation coefficient** |
| Airspace Opacity | 381 | 0.714 (0.601, 0.826) *** | 0.577 (0.506, 0.641) *** |
| Atelectasis | 296 | 0.489 (0.333, 0.645) *** | 0.348 (0.244, 0.444) *** |
| Cardiomegaly | 229 | 0.679 (0.535, 0.823) *** | 0.592 (0.501, 0.670) *** |
| Consolidation | 120 | 1.155 (0.674, 1.635) *** | 0.384 (0.220, 0.527) *** |
| Edema | 124 | 0.642 (0.459, 0.826) *** | 0.548 (0.411, 0.660) *** |
| Enlarged Cardiomediatinum | 668 | 1.974 (1.608, 2.340) *** | 0.428 (0.364, 0.488) *** |
| Lung Lesion | 50 | 0.218 (0.088, 0.349) ** | 0.509 (0.268, 0.689) *** |
| Pleural Effusion | 159 | 0.632 (0.489, 0.776) *** | 0.690 (0.599, 0.764) *** |
| Pneumothorax | 11 | 0.446 (0.108, 0.783) * | 0.734 (0.240, 0.926) * |
| Support Devices | 327 | 0.211 (0.172, 0.250) *** | 0.468 (0.378, 0.548) *** |
| All pathologies | 2365 | 0.109 (0.083, 0.135) *** | 0.285 (0.248, 0.322) *** |
| * p-value < 0.05, ** p-value < 0.01, *** p-value < 0.001 |  |  |  |

We found that for all pathologies, model confidence was positively correlated with IoU saliency method pipeline performance. The p-values for all coefficients were below 0.001 except for the coefficients for Pneumothorax (n=11) and Lung Lesion (n=50), the two pathologies for which we had the fewest positive examples. Of all the pathologies, model confidence for positive predictions of Enlarged Cardiomediastinum had the largest linear regression coefficient with IoU saliency method pipeline performance (1.974, p-value<0.001). Model confidence for positive predictions of Pneumothorax had the largest Spearman correlation coefficient with IoU saliency method pipeline performance (0.734, p-value<0.05), followed by Pleural Effusion (0.690, p-value<0.001). Combining all pathologies (n=2365), the linear regression coefficient was 0.109 (95% CI [0.083, 0.135]), and the Spearman correlation coefficient was 0.285 (95% CI [0.248, 0.322]).

We also performed analogous experiments using hit/miss as the dependent variable on the true positive slice of the test set (CXRs with both the most representative point identified by the saliency method/human benchmark and also the ground-truth segmentations) (Extended Data Fig. 9). Since every heat map contains a maximally activated point (the pixel with the highest value) regardless of model probability output, using the full dataset has limited value since false positives are due to metric set up and are not associated with model probability. We found that model confidence was positively correlated with hit/miss saliency method pipeline performance for four out of 10 pathologies.

## Discussion

The purpose of this work was to evaluate the performance of some of the most used saliency methods for deep learning explainability using a variety of model architectures. We establish the first human benchmark for CXR segmentation in a multilabel classification setup and demonstrate that saliency maps are consistently worse than expert radiologists regardless of model classification AUROC. We use qualitative and quantitative analyses to establish that saliency method localization performance is most inferior to expert localization performance when a pathology is smaller in size or has shapes that are more complex, suggesting that deep learning explainability as a clinical interface may be less reliable and less useful when used for pathologies with those characteristics. We also show that model assurance is positively correlated with saliency method localization performance, which could indicate that saliency methods are safer to use as a decision aid to clinicians when the model has made a positive prediction with high confidence.

Because ground-truth segmentations for medical imaging are time-consuming and expensive to obtain, the current norm in medical imaging—both in research and in industry—is to use classification models on which saliency methods are applied post-hoc for localization, highlighting the need for investigations into the reliability of these methods in clinical settings^46, 47^. There are public CXR datasets containing image-level labels annotated by expert radiologists (e.g., the CheXpert validation set), multilabel bounding box annotations (e.g., ChestX-ray8^48^ and VinDr-CXR^49^), and segmentations for a single pathology (e.g., SIIM-ACR Pneumothorax Segmentation^50^). To our knowledge, however, there are no other publicly available CXR datasets with multilabel pixel-level expert segmentations. By publicly releasing a development dataset, CheXlocalize, of 234 images with 643 expert segmentations, we hope to encourage the further development of saliency methods and other explainability techniques for medical imaging.

Our work has several potential implications for human-AI collaboration in the context of medical decision-making. Heat maps generated using saliency methods are advocated as clinical decision support in the hope that they not only improve clinical decision-making, but also encourage clinicians to trust model predictions^51–53^. Many of the large CXR vendors^54–56^ use localization methods to provide pathology visualization in their computer-aided detection (CAD) products. In addition to being used for clinical interpretation, saliency method heat maps are also used for the evaluation of CXR interpretation models, for quality improvement (QI) and quality assurance (QA) in clinical practices, and for dataset annotation^57^. Explainable AI is critical in high-stakes contexts such as healthcare, and saliency methods have been used successfully to develop and understand models generally. Indeed, we found that the saliency method pipeline significantly outperformed the human benchmark on two pathologies when using mIoU as an evaluation metric. However, our work also suggests that saliency methods are not yet reliable enough to validate individual clinical decisions made by a model. We found that saliency method localization performance, on balance, performed worse than expert localization across multiple analyses and across many important pathologies (our findings are consistent with recent work focused on localizing a single pathology, Pneumothorax, in CXRs^58^). We hypothesize that this could be an algorithmic artifact of saliency methods, whose relatively small heat maps (14×14 for Grad-CAM) are interpolated to the original image dimensions (usually 2000×2000), resulting in coarse resolutions. If used in clinical practice, heat maps that incorrectly highlight medical images may exacerbate well-documented biases (chiefly, automation bias) and erode trust in model predictions (even when model output is correct), limiting clinical translation^22^.

Since IoU computes the overlap of two segmentations but pointing game hit rate better captures diagnostic attention, we suggest using both metrics when evaluating localization performance in the context of medical imaging. While IoU is a commonly used metric for evaluating semantic segmentation outputs, there are inherent limitations to the metric in the pathological context. This is indicated by our finding that even the human benchmark segmentations had low overlap with the ground-truth segmentations (the highest expert mIoU was 0.720 for Cardiomegaly). One potential explanation for this consistent underperformance is that pathologies can be hard to distinguish, especially without clinical context. Furthermore, whereas many people might agree on how to segment, say, a cat or a stop sign in traditional computer vision tasks, radiologists use a certain amount of clinical discretion when defining the boundaries of a pathology on a CXR. There can also be institutional and geographic differences in how radiologists are taught to recognize pathologies, and studies have shown that there can be high interobserver variability in the interpretation of CXRs^59–61^. We sought to address this with the hit/miss evaluation metric, which highlights when two radiologists share the same diagnostic intention, even if it is less exact than IoU in comparing segmentations directly. The human benchmark localization using hit rate was above 0.9 for four pathologies (Pneumothorax, Cardiomegaly, Enlarged Cardiomediastinum, and Support Devices); these are pathologies for which there is often little disagreement between radiologists about where the pathologies are located, even if the expert segmentations are noisy. Further work is needed to demonstrate which segmentation evaluation metrics, even beyond IoU and hit/miss, are more appropriate for certain pathologies and downstream tasks when evaluating saliency methods for the clinical setting.

Our work builds upon several studies investigating the validity of saliency maps for localization^62–64^ and upon some early work on the trustworthiness of saliency methods to explain DNNs in medical imaging^47^. However, as recent work has shown^32^, evaluating saliency methods is inherently difficult given that they are post-hoc techniques. To illustrate this, consider the following models and saliency methods as described by some oracle: (1) a model *M_bad* that has perfect AUROC for a given image classification task, but that we know does *not* localize well (i.e. because the model picks up on confounders in the image); (2) a model *M_good* that also has perfect AUROC, but that we know *does* localize well (i.e. is looking at relevant regions of the image); (3) a saliency method *S_bad* that does *not* properly reflect the model’s attention; and (4) a saliency method *S_good* that *does* properly reflect the model’s attention. Let us say that we are evaluating the following pipeline: we first classify an image and we then apply a saliency method post hoc. Imagine that our evaluation reveals poor localization performance as measured by mIoU or hit rate (as was the case in our findings). There are three possible pipelines (combinations of model and saliency method) that would lead to this scenario: (1) *M_bad* + *S_good*; (2) *M_good* + *S_bad*; and (3) *M_bad* + *S_bad*. The first scenario (*M_bad + S_good*) is the one for which saliency methods were originally intended: we have a working saliency method that properly alerts us to models picking up on confounders. The second scenario (*M_good + S_bad*) is our nightmare scenario: we have a working model whose attention is appropriately directed, but we reject it based on a poorly localizing saliency method. Because all three scenarios result in poor localization performance, it is difficult—if not impossible—to know whether poor localization performance is attributable to the model or to the saliency method (or to both). While we cannot say whether models or saliency methods are failing in the context of medical imaging, we can say that we should not rely on saliency methods to evaluate model localization. Future work should explore potential techniques for localization performance attribution.

There are several limitations of our work. First, we did not investigate the impact of pathology prevalence in the training data on saliency method localization performance. Second, some pathologies, such as effusions and cardiomegaly, are in similar locations across frontal view CXRs, while others, such as lesions and opacities, can vary in locations across CXRs. Future work could investigate how the location of pathologies on a CXR in the training/test data distribution, and the consistency of those locations, affect saliency method localization performance. Third, while we compared saliency method-generated pixel-level segmentations to human expert pixel-level segmentations, future work might explore how saliency method localization performance changes when comparing bounding-box annotations, instead of pixel-level segmentations. Fourth, we explored post-hoc interpretability methods given their prevalence in the context of medical imaging, but we hope that by publicly releasing our development dataset of pixel-level expert segmentations we can facilitate the development of models that make use of ground-truth segmentations during training^57^. Fifth, the lack of a given finding can in certain cases inform clinical diagnoses. A common example of this is the lack of normal lung tissue pattern towards the edges of the thoracic cage, which is used to detect pneumothorax. For any characteristic pattern, both the absence and the presence provide diagnostic information to the radiologist. For example, the absence of a pleural effusion pattern is also used to rule out pleural effusion. For any characteristic radiological pattern, both the presence as well as the absence contribute to the final radiology report. Future work can explore counterfactual visual explanations that are similar to the counterfactual diagnostic process of a radiologist. Sixth, future work should further explore the potentially confounding effect of model calibration on the evaluation of saliency methods, especially when using segmentation, as opposed to classification, models. Finally, the impact of saliency methods on the trust and efficacy of users is underexplored.

In conclusion, we present a rigorous evaluation of a range of saliency methods and a dataset of pixel-level expert segmentations, which can serve as a foundation for future work exploring deep learning explainability techniques. This work is a reminder that care should be taken when leveraging common saliency methods to validate individual clinical decisions in deep learning-based workflows for medical imaging.

## Methods

### Ethical and information governance approvals

A formal Stanford IRB review was conducted for the original collection of the CheXpert dataset. The IRB waived the requirement to obtain informed consent as the data were retrospectively collected and fully anonymized.

### Dataset and clinical taxonomy

#### Dataset description

The localization experiments were performed using CheXpert, a large public dataset for chest X-ray interpretation. The CheXpert dataset contains 224,316 chest X-rays for 65,240 patients labeled for the presence of 14 observations (13 pathologies and an observation of “No Finding”) as positive, negative, or uncertain. The CheXpert validation set consists of 234 chest X-rays from 200 patients randomly sampled from the full dataset and was labeled according to the consensus of three board-certified radiologists. The test set consists of 668 chest X-rays from 500 patients not included in the training or validation sets and was labeled according to the consensus of five board-certified radiologists. See Extended Data Fig. 10 for test set summary statistics. “Lung Opacity” in the CheXpert dataset is the equivalent of “Airspace Opacity” in the CheXlocalize dataset.

#### Ground-truth segmentations

The chest X-rays in our validation set and test set were manually segmented by two board-certified radiologists with 18 and 27 years of experience, using the annotation software tool MD.ai^65^ (see Supplementary Figs. S12 through S14). The radiologists were asked to contour the region of interest for all observations in the chest X-rays for which there was a positive ground-truth label in the CheXpert dataset. For a pathology with multiple instances, all the instances were contoured. For Support Devices, radiologists were asked to contour any implanted or invasive devices (including pacemakers, PICC/central catheters, chest tubes, endotracheal tubes, feeding tubes and stents), and to ignore ECG lead wires or external stickers visible in the chest X-ray.

#### Benchmark segmentations

To evaluate expert performance on the test set using IoU, three radiologists, certified in Vietnam with 9, 10, and 18 years of experience, were asked to segment the regions of interest for all observations in the chest X-rays for which there was a positive ground-truth label in the CheXpert dataset. These radiologists were also provided the same instructions for contouring as were provided to the radiologists drawing the ground-truth segmentations. To extract the “maximally activated” point from the benchmark segmentations, we asked the same radiologists to locate each pathology present on each CXR using only a single most representative point for that pathology on the CXR (see Supplementary Figs. S1 through S11 for the detailed instructions given to the radiologists). There was no overlap between these three radiologists and the two who drew the ground-truth segmentations.

### Classification network architecture and training protocol

#### Multi-label classification model

The model takes as input a single-view chest X-ray and outputs a probability for each of the 14 observations. If more than one view is available, the model outputs the maximum probability of the observations across the views. Each chest X-ray was resized to 320×320 pixels and normalized before it was fed into the network. We used the same image resolutions as CheXpert^36^ and CheXNet^2^, which demonstrated radiologist-level performance on external test sets with 320×320 images. There are models that are commercially deployed and have similar dimensions. For example, the architecture used by medical AI software vendor Annalise.ai^66^ is based on EfficientNet^67^, which takes input of 224×224. Chest X-rays were normalized prior to being fed into the network by subtracting the mean of all images in the CheXpert training set and then dividing by the standard deviation of all images in the CheXpert training set. The model architectures DenseNet121, ResNet152, and Inception-v4 were used. Cross-entropy loss was used to train the model. The Adam optimizer^68^ was used with default β-parameters of β1 = 0.9 and β2 = 0.999. The learning rate was tuned for the different model architectures using grid search (over learning rates of 1e-3, 1e-4, and 1e-5). The best learning rate for each architecture was: 1e-4 for DenseNet121, 1e-5 for ResNet152, and 1e-5 for Inceptionv4. Batches were sampled using a fixed batch size of 16 images.

#### Ensembling

We use an ensemble of checkpoints to create both predictions and saliency maps to maximize model performance. In order to capture uncertainties inherent in radiograph interpretation, we train our models using four uncertainty handling strategies outlined in CheXpert: Ignoring, Zeroes, Ones, and 3-Class Classification. For each of the four uncertainty handling strategies, we train our model three separate times, each time saving the 10 checkpoints across the three epochs with the highest average AUC across 5 observations selected for their clinical importance and prevalence in the validation set: Atelectasis, Cardiomegaly, Consolidation, Edema, and Pleural Effusion. In total, after training, we have saved 4 x 30 = 120 checkpoints for a given model. Then, from the 120 saved checkpoints for that model, we select the top 10 performing checkpoints for each pathology. For each CXR and each task, we compute the predictions and saliency maps using the relevant checkpoints. We then take the mean both of the predictions and of the saliency maps to create the final set of predictions and saliency maps for the ensemble model. See Supplementary Table S1 for the performance of each model architecture (DenseNet121, ResNet152, and Inception-v4) on each of the pathologies.

#### Evaluating localization performance

Saliency methods were used to visualize the decision made by the classification network. Each saliency map was resized to the original image dimension using bilinear interpolation. It was then normalized using min-max normalization and converted into a binary segmentation using binary thresholding (Otsu’s method). For Occlusion, we used a window size of 40 and a stride of 40 for each CXR.

Localization performance of each segmentation was evaluated using Intersection over Union (IoU) score. The IoU is the ratio between the area of overlap and the area of union between the ground-truth and the predicted segmentations, ranging from 0 to 1 (0 signifies no overlap and 1 signifies perfectly overlapping segmentations). We report the mean Intersection over Union (mIoU) over 1,000 bootstrap replicates on the test set, along with the 95% confidence intervals using the 2.5^th^ and 97.5^th^ percentiles of the empirical distribution.

For the evaluation of DenseNet121+Integrated Gradients using IoU, we applied box filtering of kernel size 100 to smooth the pixelated map. For the evaluation of ResNet152+Integrated Gradients and of Inception-v4+Integrated Gradients using IoU, we applied box filtering of kernel size 50. For the evaluation of DeepLIFT using IoU, we applied box filtering of kernel size 50. For the evaluation of LRP using IoU, we applied box filtering of kernel size 80. The kernel sizes were tuned on the validation set. The noisy map is not a concern for hit rate because a single max pixel is extracted for the entire image.

In Extended Data Fig. 1, we report mIoU localization performance using different saliency map thresholding values. We first applied min-max normalizations to the saliency maps so that each value gets transformed into a decimal between 0 and 1. We then passed in a range of threshold values from 0.2 to 0.8 to create binary segmentations and calculated the mIoU score per pathology under each threshold on the validation set.

In Extended Data Fig. 2, we report the precision, recall/sensitivity, and specificity values of the saliency method pipeline and the human benchmark segmentations on the test set.

For this, we treat each pixel in the saliency method pipeline and the human benchmark segmentations as a classification, use each pixel in the ground-truth segmentation as the ground-truth label, and calculate precision, recall/sensitivity, and specificity over all CXRs for each pathology. Precision is defined as the number of total TP pixels / (number of total TP + FP pixels). Recall is defined as the number of total TP pixels / (number of total TP + FN pixels). Specificity is defined as the number of total TN pixels / (number of total TN + FP pixels).

In Extended Data Fig. 4, we report the saliency method pipeline test set localization performance on the full dataset using mIoU. For this, we ensure that the final binary segmentation is consistent with model probability output by applying another layer of thresholding such that the segmentation mask produced all zeros if the predicted probability was below a chosen level. The probability threshold is searched on the interval of [0,0.8] with steps of 0.1. The exact value is determined per pathology by maximizing the mIoU on the validation set.

In Extended Data Figs. 5 and 6, we report the percentage decrease from human benchmark localization performance to saliency method pipeline localization performance on the test set. To obtain the 95% confidence interval per pathology on the percentage decrease from human benchmark localization performance to saliency method pipeline localization performance, we first extracted the percentage decrease statistic ([human benchmark mIoU or hit rate-saliency method pipeline mIoU or hit rate] / human benchmark mIoU or hit rate x 100) from each of the 1,000 human benchmark and the 1,000 saliency method pipeline mIoU/hit rate bootstrap replicates for each pathology. In doing so, we created the bootstrap distribution of the percentage decrease statistic. We reported the 95% CI using the 2.5^th^ and 97.5^th^ percentiles of the empirical distribution. To obtain the 95% CI on the average percentage decrease over all pathologies, the methodology is the same: we created bootstrap replicates of the average human benchmark and saliency method pipeline mIoUs/hit rates over all pathologies, extracted the percentage decrease statistic from each replicate, and then reported the 95% CI using the 2.5^th^ and 97.5^th^ percentiles of the empirical distribution.

### Statistical analyses

#### Pathology Characteristics

The pathology characteristics used in all regressions were calculated on the ground-truth annotations. The four characteristics are defined as follows: (1) Number of instances is the number of separate segmentations drawn by the radiologist for a given pathology. (2) Size is the area of the pathology divided by the total image area. (3) and (4) Elongation and irrectangularity are geometric features that measure shape complexities. They were designed to quantify what radiologists qualitatively describe as focal or diffused. To calculate the metrics, a rectangle of minimum area enclosing the contour is fitted to each pathology. Elongation is defined as the ratio of the rectangle’s longer side to shorter side. Irrectangularity = 1 - (area of segmentation/area of enclosing rectangle), with values ranging from 0 to 1 (1 being very irrectangular). When there were multiple instances within one pathology, we used the characteristics of the dominant instance (largest in perimeter). All geometric features are normalized using min-max normalization per-pathology before aggregation so that they are comparable on scales of magnitudes.

#### Model Confidence

We used the probability output of the DNN architecture for model confidence. The probabilities were on the similar scale of 0-1 and we did not apply min-max normalization. We report the 95% confidence interval and p-value of the regression coefficients using Student’s t-distribution.

For the statistical analyses on the *full* dataset to determine whether there was any correlation between the model’s confidence in its prediction and saliency method pipeline performance using IoU (Table 3), we ensure that the final binary segmentation is consistent with model probability output by applying another layer of thresholding such that the segmentation mask produced all zeros if the predicted probability was below a chosen level. The probability threshold is searched on the interval of [0,0.8] with steps of 0.1. The exact value is determined per pathology by maximizing the mIoU on the validation set.

For the statistical analyses to determine whether there was any correlation between the model’s confidence in its prediction and saliency method pipeline performance using hit/miss (Extended Data Fig. 9), we used the true positive slice of the dataset (CXRs with both the most representative point identified by the saliency method/human benchmark and also the ground-truth segmentations). Since every heat map contains a maximally activated point (the pixel with the highest value) regardless of model probability output, using the full dataset has limited value since false positives are due to metric set up and are not associated with model probability.

## Data Availability

The CheXlocalize dataset is available here: https://stanfordaimi.azurewebsites.net/datasets/abfb76e5-70d5-4315-badc-c94dd82e3d6d. The CheXpert dataset is available here https://stanfordmlgroup.github.io/competitions/chexpert/.

## Code Availability

The code used to produce our results is available in the following public repository under the MIT License: https://github.com/rajpurkarlab/cheXlocalize.

## Data Availability

CheXpert data is available at https://stanfordmlgroup.github.io/competitions/chexpert/. The validation set and corresponding benchmark radiologist annotations will be available online for the purpose of extending the study.

## Acknowledgements

We would like to acknowledge MD.ai for generously providing us access to their annotation platform. We would like to acknowledge Weights & Biases for generously providing us access to their experiment tracking tools.

## Author Contributions

Conceptualization: P.R. and A.P. Design: P.R., A.P., A.S., X.G. and A.A. Data analysis and interpretation: A.S., X.G., A.A., P.R., A.P., S.T., C.N., V.N., J.S., and F.B. Drafting of the manuscript: A.S., X.G., A.A., and P.R. Critical revision of the manuscript for important intellectual content: A.P, S.T., C.N., V.N., J.S., F.B, A.N., and M.L. Supervision: A.N., M.L., and P.R. Research was primarily performed while A.S. was at Stanford University. M.L. and P.R. contributed equally.

## Extended Data

**Extended Data Fig. 1.**
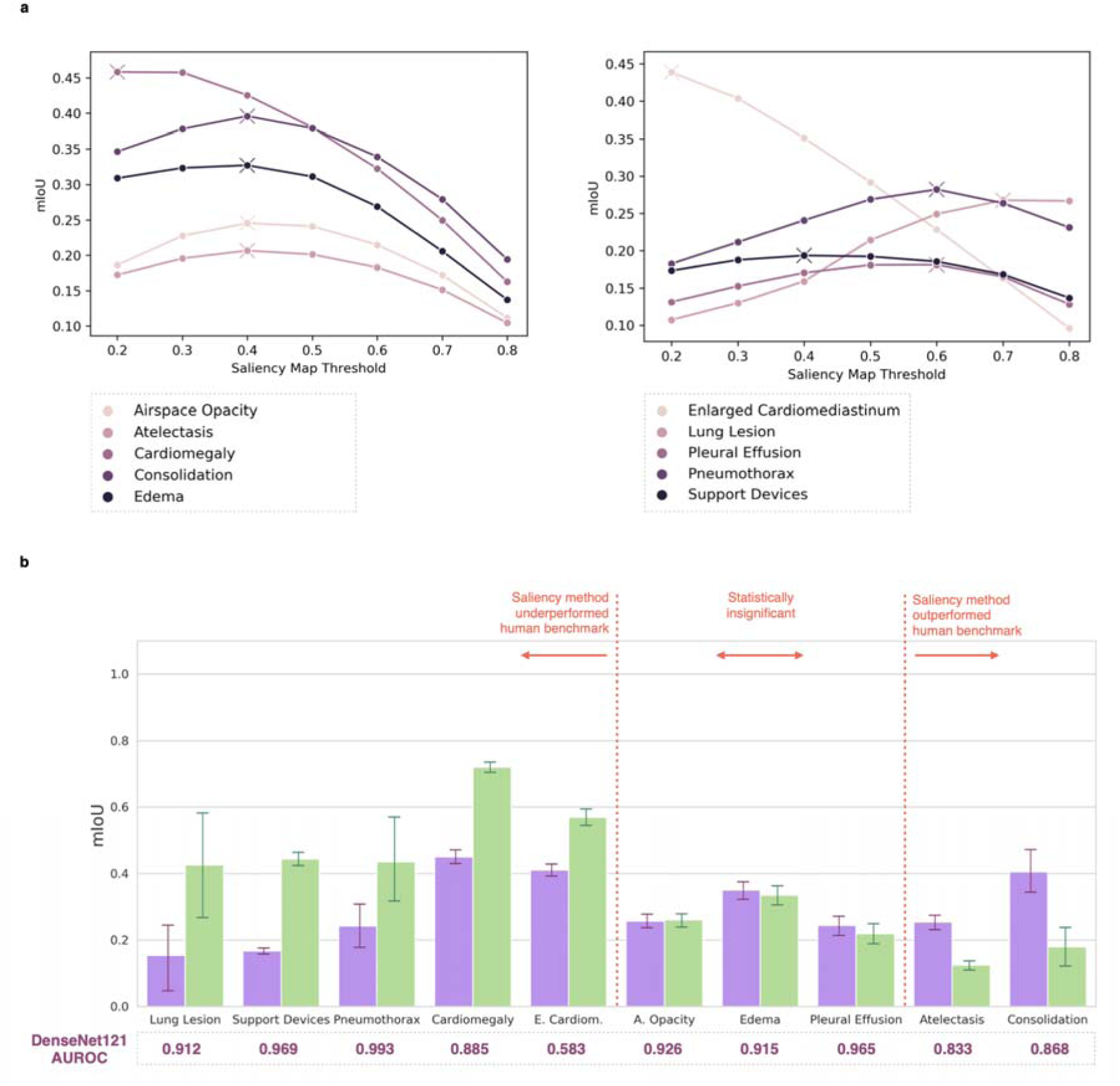
mIoU localization performance of the saliency method pipeline on the test set using threshold values tuned on the validation set. **a**, We first applied min-max normalizations to the Grad-CAM saliency maps so that each value gets transformed into a decimal between 0 and 1. We then passed in a range of threshold values from 0.2 to 0.8 to create binary segmentations and plotted the mIoU score per pathology under each threshold on the validation set. The threshold that gives the max mIoU for each pathology is marked with an “X”. Pathologies are sorted alphabetically and shown in two plots for readability. **b**, Comparing mIoU localization performances of the saliency method pipeline on the test set (using the best thresholds tuned on the validation set) and the human benchmark. We found that the saliency method pipeline outperformed the human benchmark on two pathologies and underperformed the human benchmark on five pathologies. For the remaining three pathologies, the performance differences were not statistically significant. This finding is consistent with what we report in the manuscript using Otsu’s method.

**Extended Data Fig. 2.**
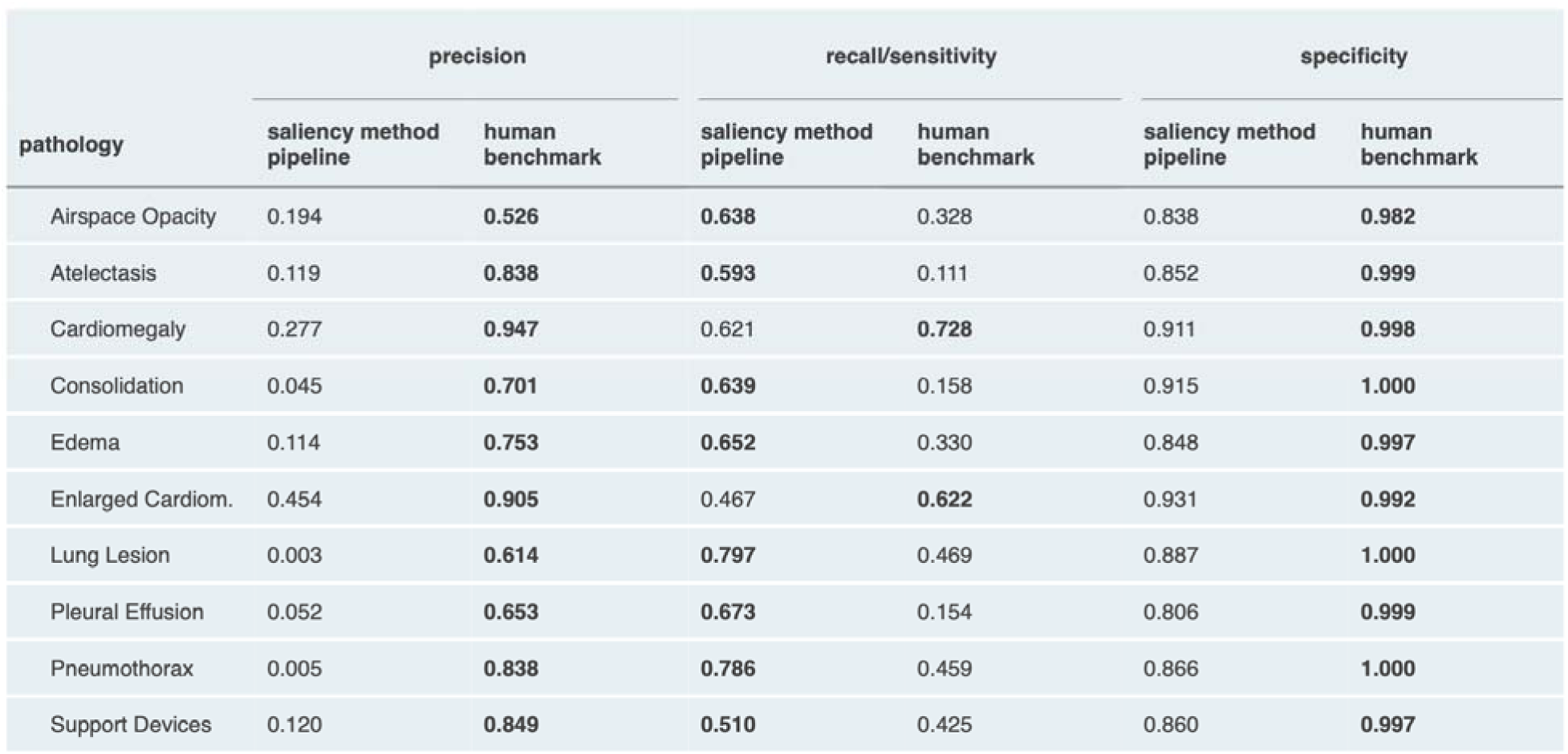
Precision, recall/sensitivity, and specificity values of the saliency method pipeline and the human benchmark segmentations on the test set. We treated each pixel in the saliency method pipeline and the human benchmark segmentations as a classification, used each pixel in the ground-truth segmentation as the ground-truth label, and calculated the precision, recall/sensitivity, and specificity over all CXRs for each pathology. For each pathology and each metric, we highlight the higher of the two (saliency method pipeline or human benchmark) in **bold**.

**Extended Data Fig. 3.**
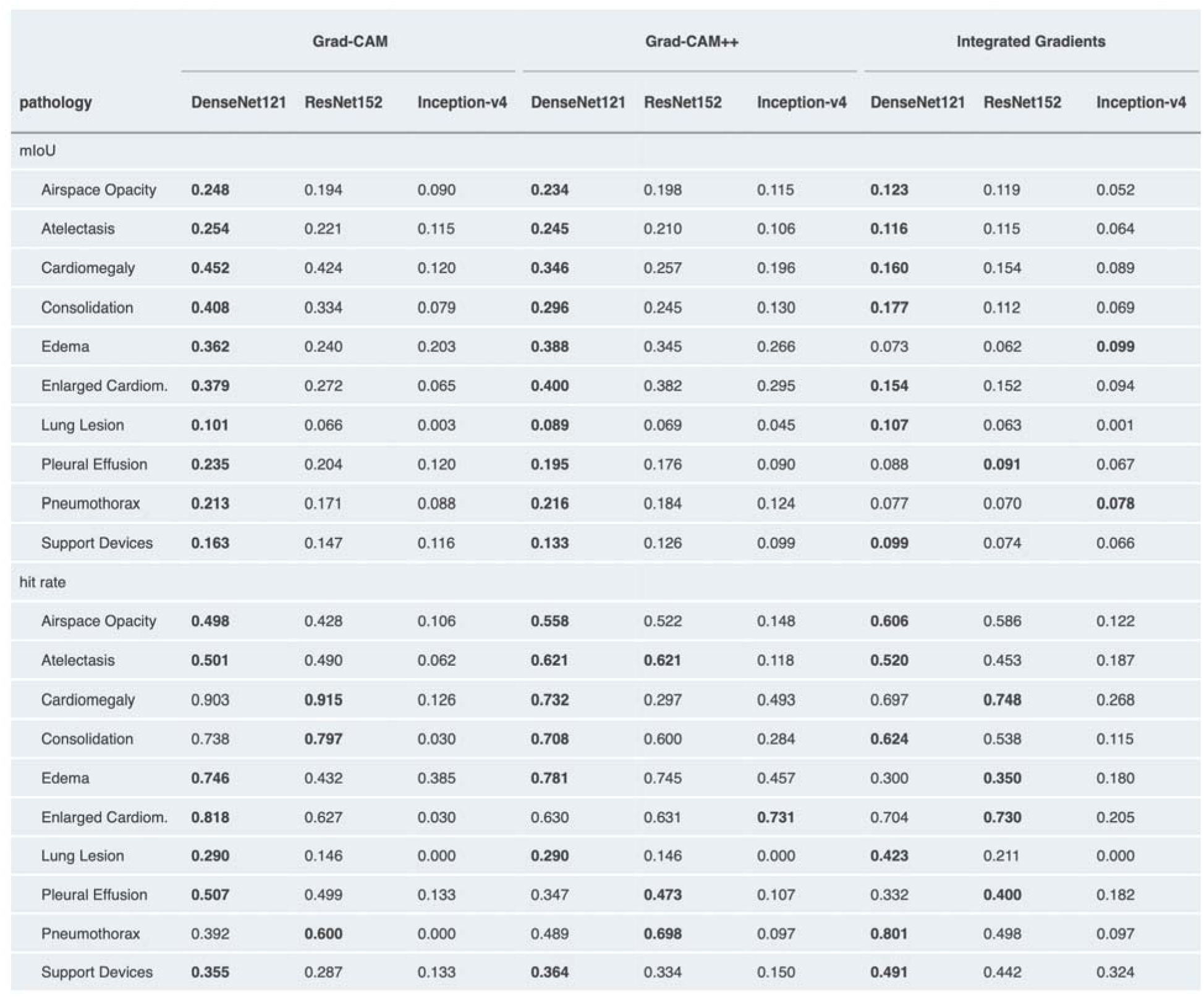
Test set localization performance for each combination of saliency method and CNN architecture. For each pathology and saliency method, we highlight the highest performing CNN architecture in **bold**.

**Extended Data Fig. 4.**
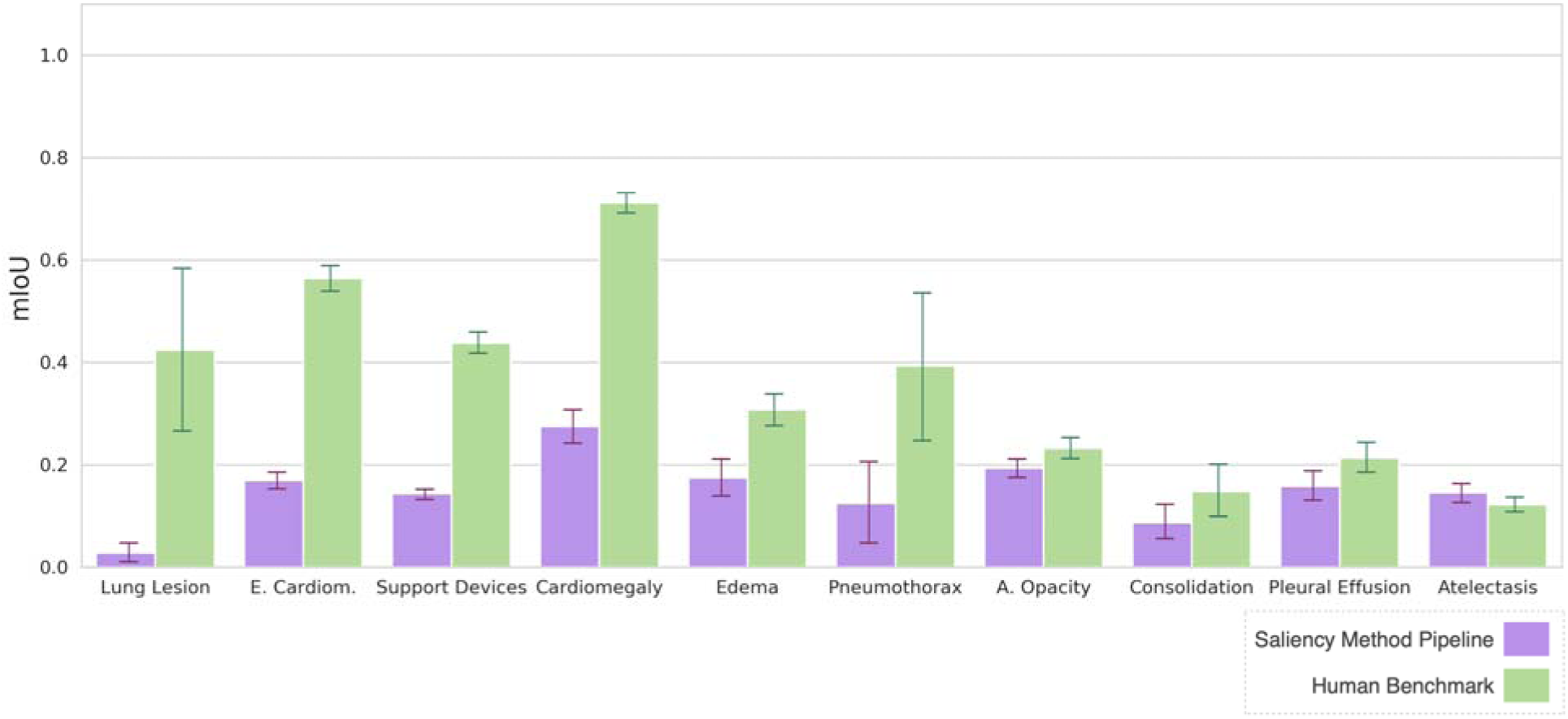
Saliency method pipeline test set localization performance on the full dataset using mIoU. True negatives (CXRs whose ground-truth label is negative for a given pathology and for which there were neither human benchmark nor saliency method pipeline segmentations for that pathology) were excluded from the metric calculation. To control for false positives, we ensure that the final binary segmentation is consistent with model probability output by applying another layer of thresholding such that the segmentation mask produced all zeros if the predicted probability was below a chosen level. The probability threshold is searched on the interval of [0,0.8] with steps of 0.1. The exact value is determined per pathology by maximizing the mIoU on the validation set. We found that on the full dataset, for seven of the 10 pathologies, the saliency method pipeline had a significantly lower mIoU than the human benchmark.

**Extended Data Fig. 5.**
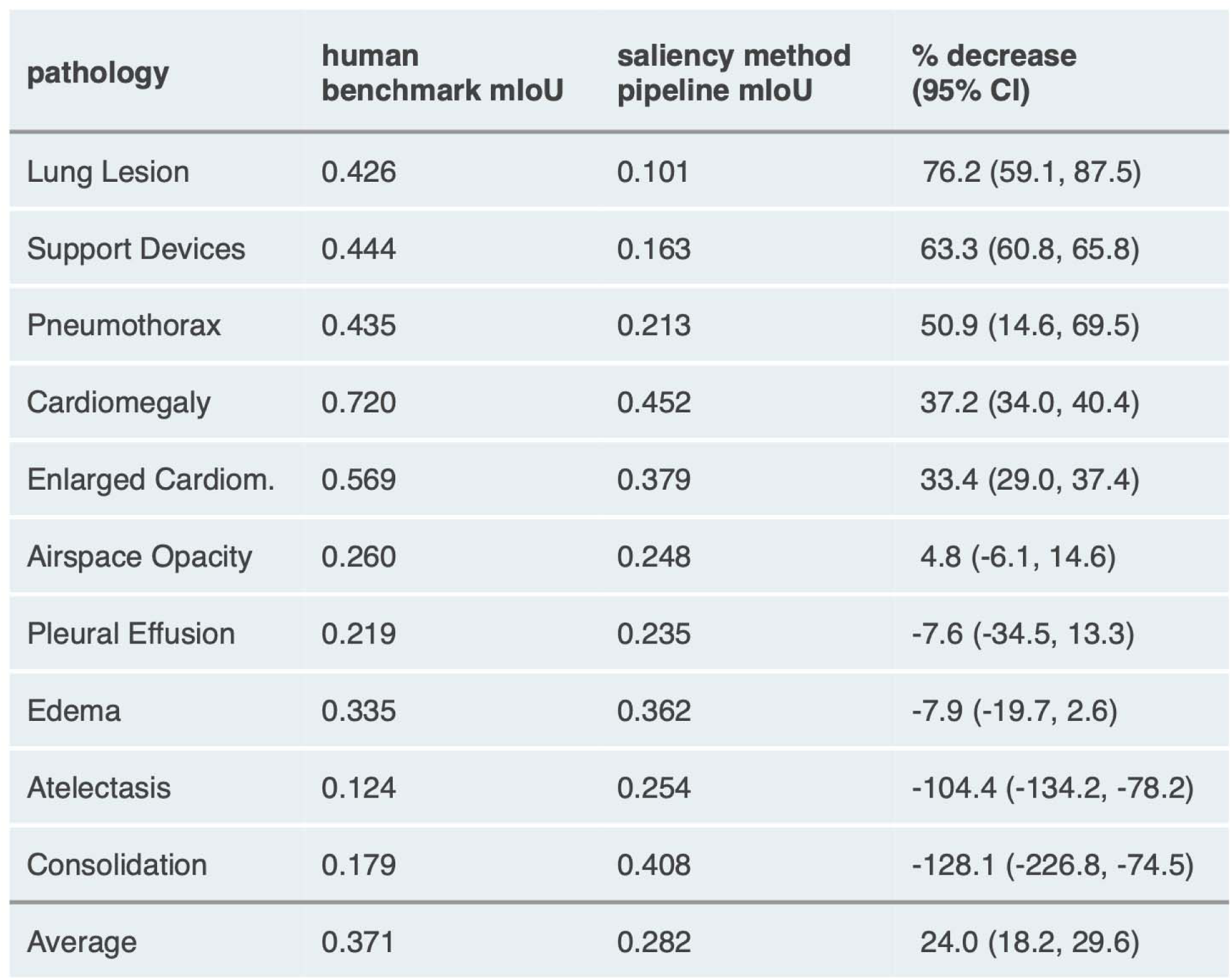
Percentage decrease from human benchmark mIoU to saliency method pipeline mIoU on the test set. Pathologies are sorted first by statistical significance of percentage decrease from human benchmark mIoU to saliency method pipeline mIoU (high to low), and then by percentage decrease from human benchmark mIoU to saliency method pipeline mIoU (high to low). We use 95% bootstrap confidence interval.

**Extended Data Fig. 6.**
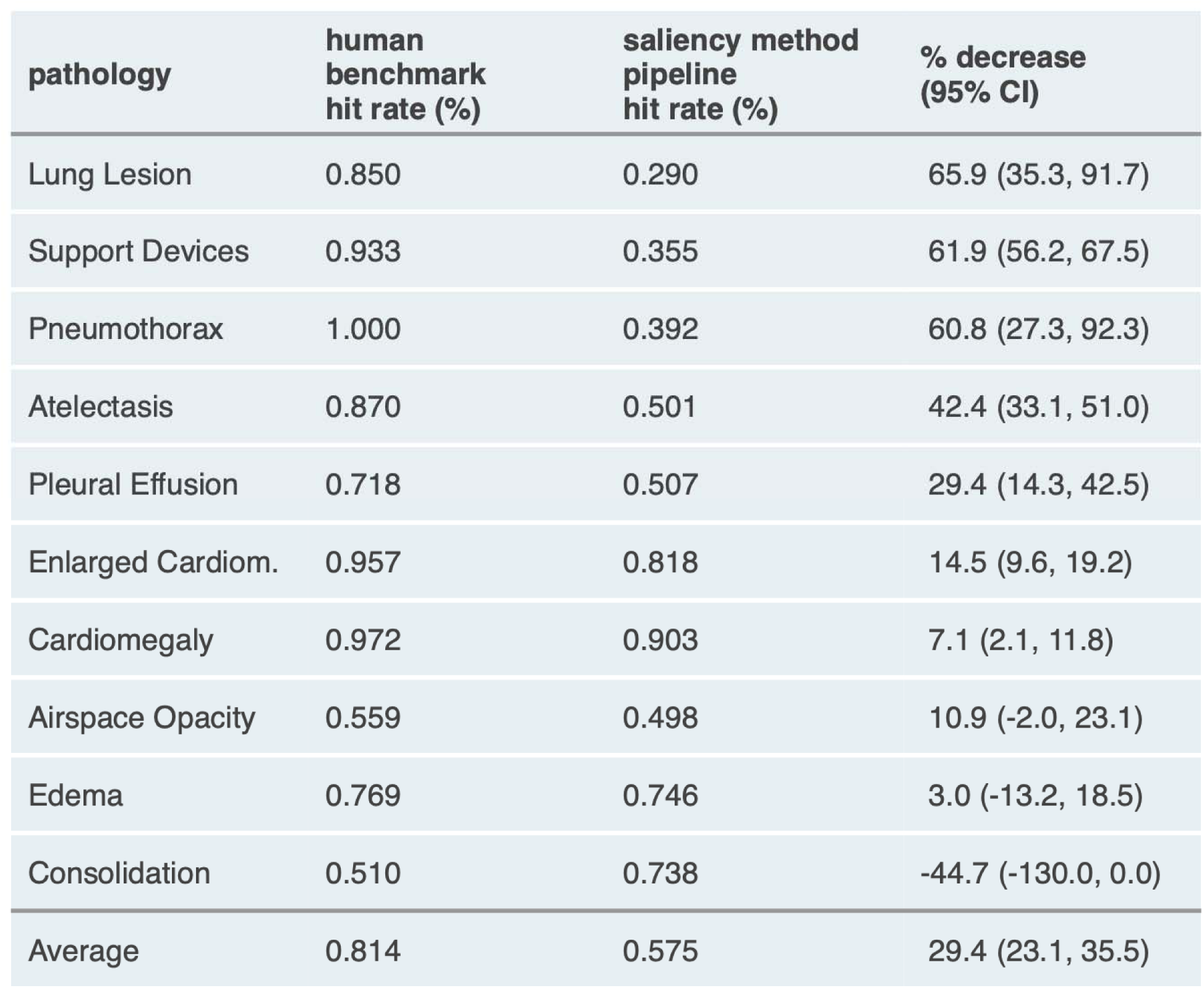
Percentage decrease from human benchmark hit rate to saliency method pipeline hit rate on the test set. Pathologies are sorted first by statistical significance of percentage decrease from human benchmark hit rate to saliency method pipeline hit rate (high to low), and then by percentage decrease from human benchmark hit rate to saliency method pipeline hit rate (high to low). We use 95% bootstrap confidence interval.

**Extended Data Fig. 7.**
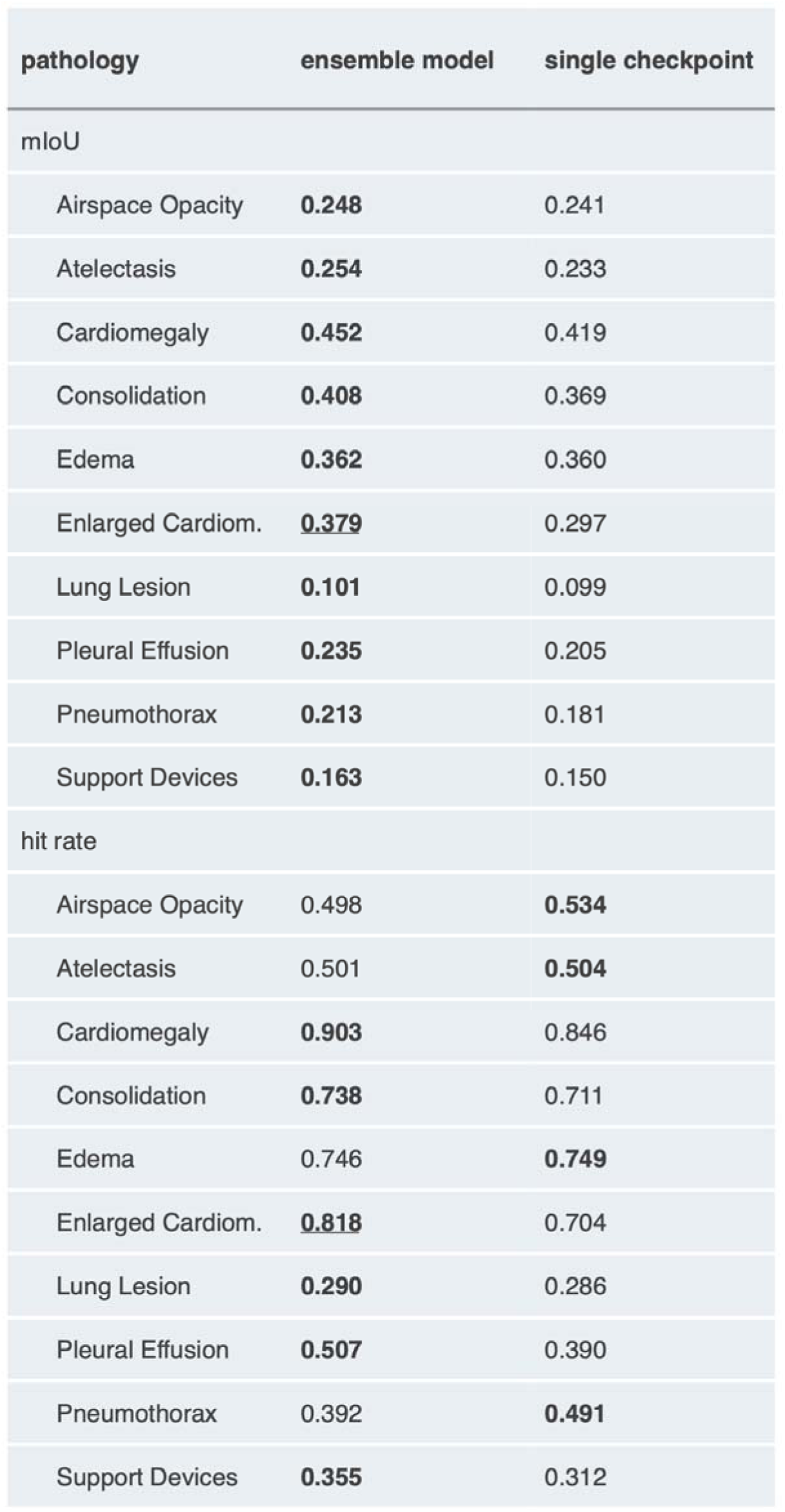
Test set saliency method pipeline localization performance using an ensemble model vs. using the top performing single checkpoint for each pathology. For each pathology, we highlight in **bold** the model (ensemble or single checkpoint) that has the higher metric, and we <u>underline</u> it if the difference is statistically significant (using 95% bootstrap confidence interval).

**Extended Data Fig. 8.**
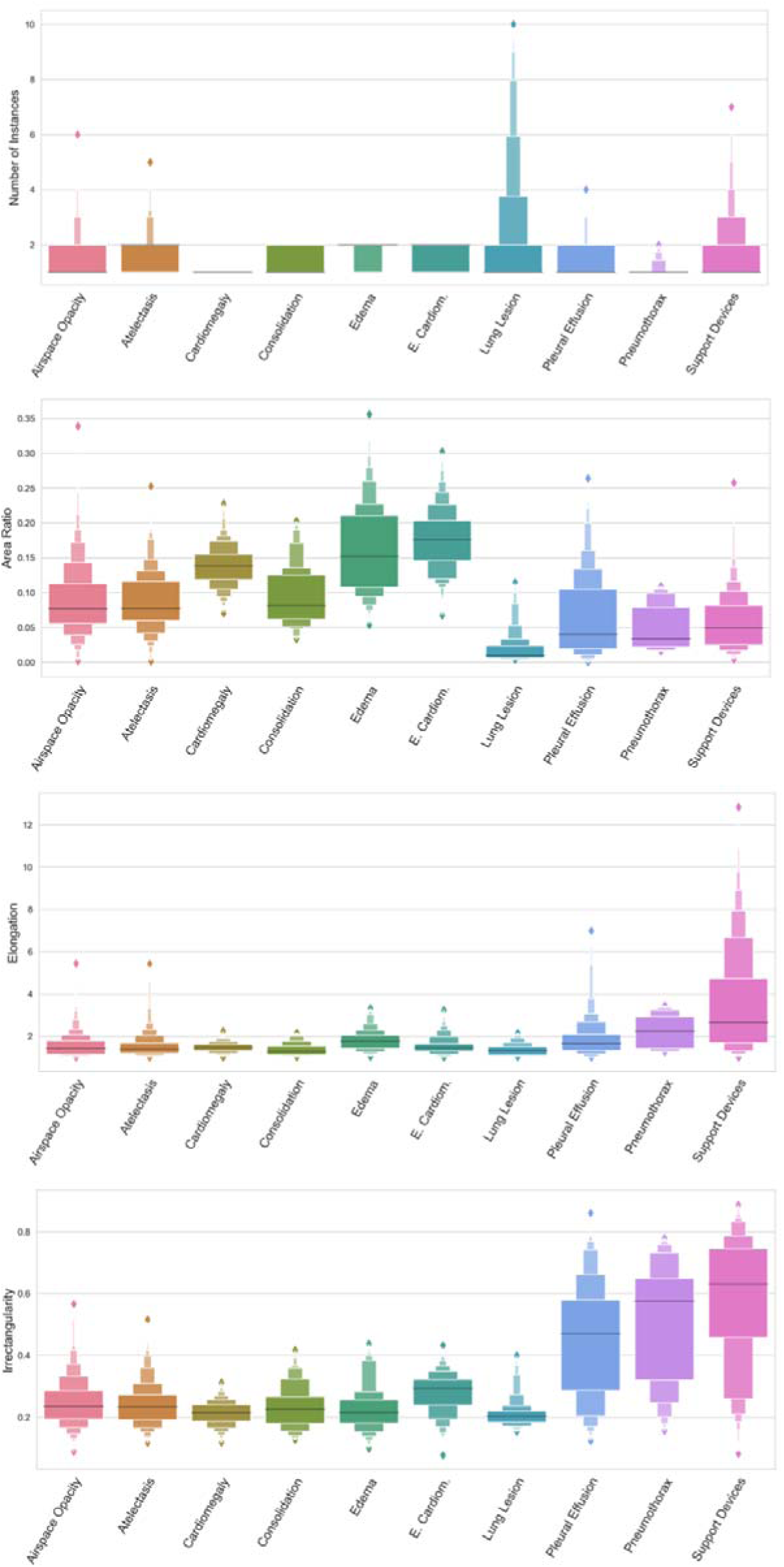
Test set distribution of four geometric features across all 10 pathologies. The black horizontal line in each box indicates the median feature value for that pathology, and each successive level outward contains half of the remaining data. The height of the box indicates the range of feature values in the quantile.

**Extended Data Fig. 9.**
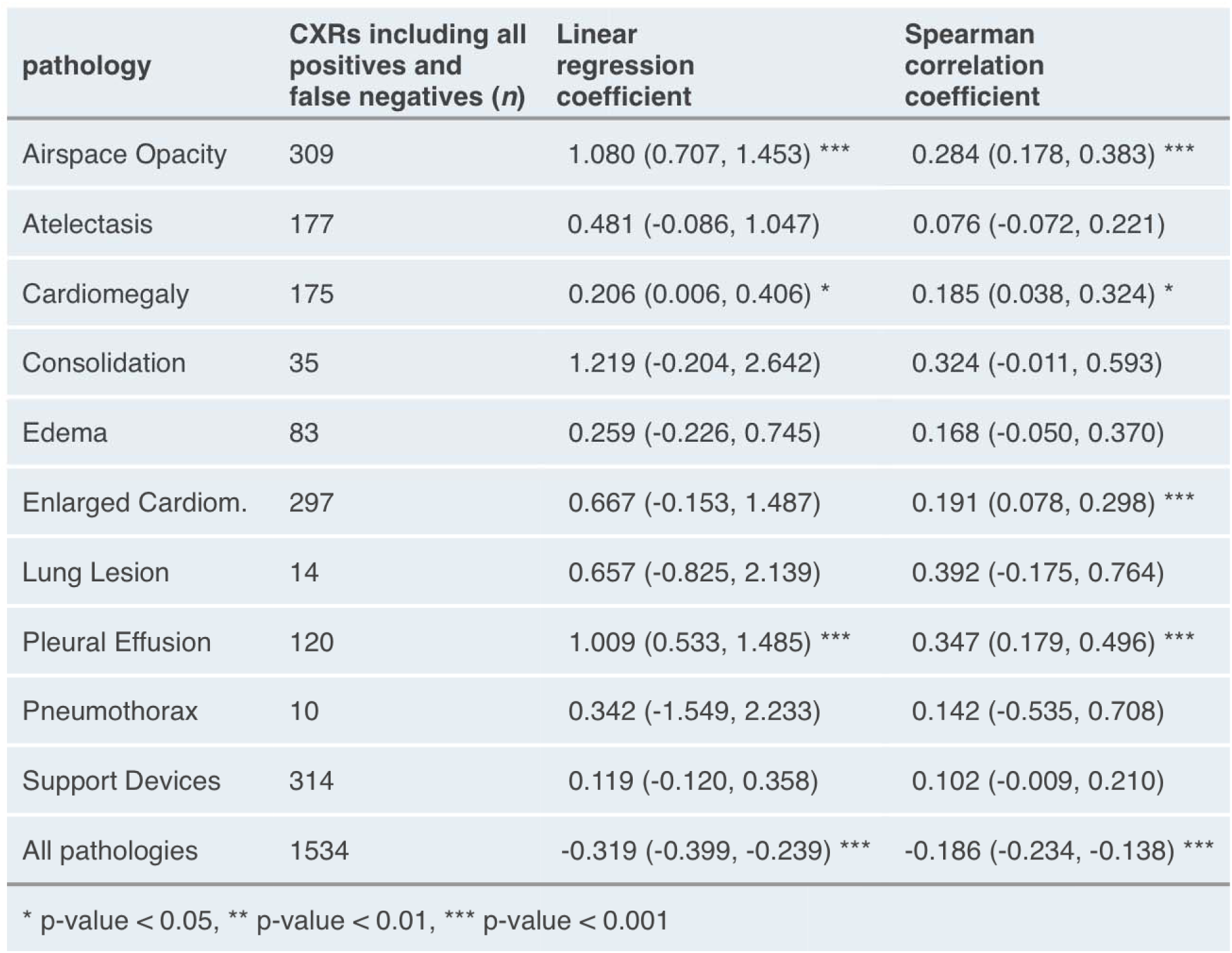
Hit/miss: Coefficients from regressions on model assurance. Statistical analysis to determine whether there was any correlation between the model’s confidence in its prediction and saliency method pipeline performance using hit/miss. We used the true positive slice of the dataset (CXRs with both the most representative point identified by the saliency method/human benchmark and also the ground-truth segmentations). Since every heat map contains a maximally activated point (the pixel with the highest value) regardless of model probability output, using the full dataset has limited value since false positives are due to metric set up and are not associated with model probability.

**Extended Data Fig. 10.**
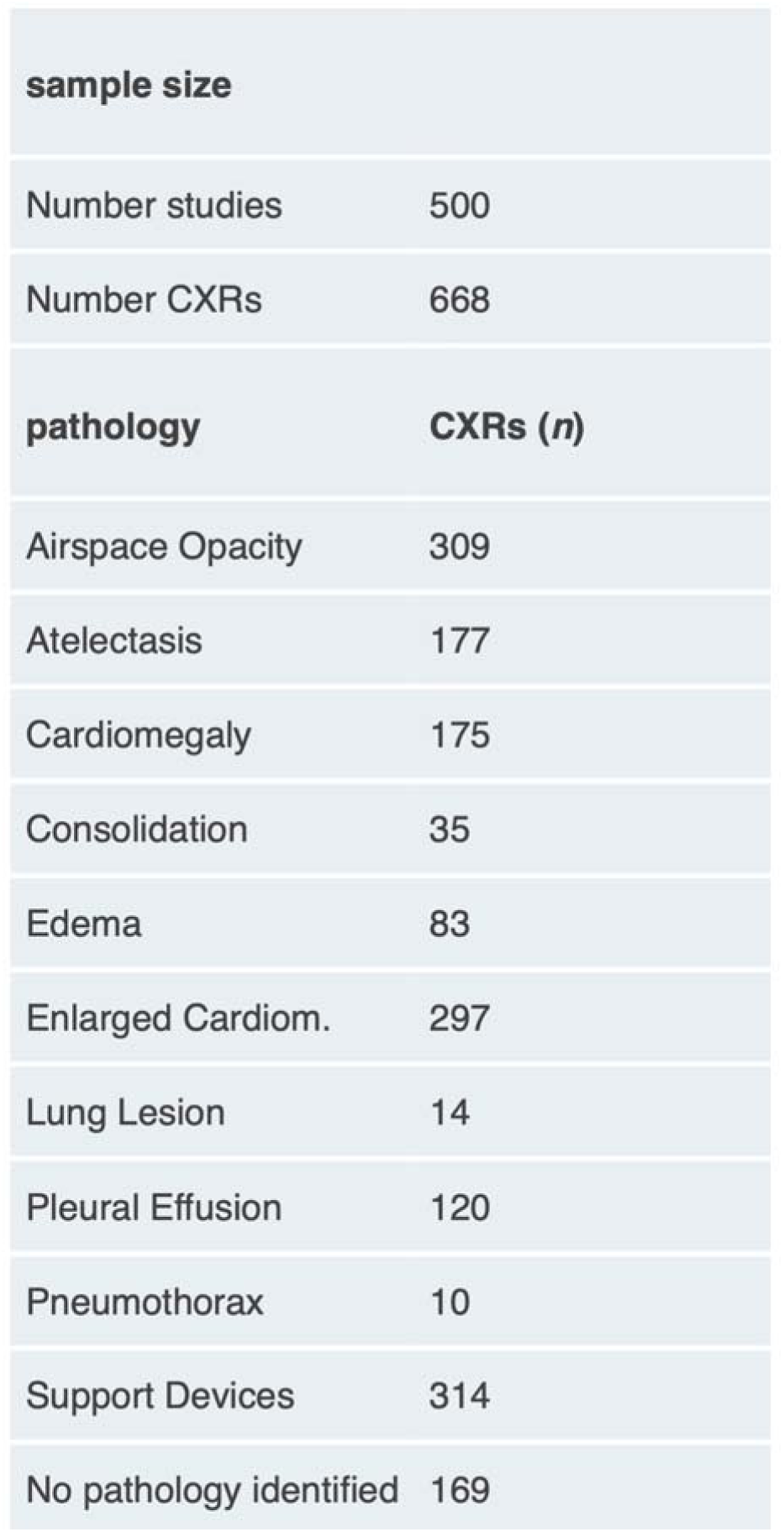
Test set summary statistics.

