## Supplemental data for "Benchmarking saliency methods for chest X-ray interpretation"

We would like to localize the following **10 observations** on a set of chest X-rays (CXRs): Enlarged Cardiomediastinum, Cardiomegaly, Lung Opacity, Lung Lesion, Edema, Consolidation, Atelectasis, Pneumothorax, Pleural Effusion, and Support Devices.

You will be given the same set of CXRs on which you earlier drew segmentations for the above 10 observations. As a reminder, each CXR has 10 ground truth labels (0 or 1) for each of the above 10 observations, and you drew segmentations on each of the assigned CXRs only for the observations that were present as determined by the ground truth labels.

Now, for these same CXRs on which you drew the segmentations, we would like for you to select **the single most salient point** on the CXR for each positive observation. Which point is most salient will depend on the observation at hand, but the point will always lie inside the segmentation you drew. For example, for Pneumothorax, this point might be wherever the pathology is most pronounced. As another example, for Cardiomegaly, the most salient point will be the center of the heart. Below, we have included descriptions and examples of what we are expecting the most salient point to be for each of the 10 observations.

For some CXRs, there are multiple instances/segmentations for a given observation (for example, if there are multiple support devices, or if the patient has bilateral pleural effusion). Even when there are multiple segmentations, please only select **one single point** for that observation on the CXR; this point will necessarily lie inside *one* of the segmentations that you drew for that observation.

At the end of this exercise, there should be one point for each positive label for each CXR. For example, if a CXR has positive labels for Cardiomegaly, Lung Opacity, and Edema, then there should be three series of segmentations with three points for that CXR.

As before, we understand that you may not always agree with the ground truth labels. However, we ask that you please attempt to annotate all of the positive observations according to the ground truth labels. Also, please note that you do *not* need to select a point if the CXR has a positive label for No Finding (however, a CXR with a No Finding label may have a positive label for Support Devices, which does require a point!).

**Supplementary Fig. S1 | General instructions given to benchmark radiologists on how to select the most representative point on a CXR.**

### Enlarged Cardiomediastinum

Enlarged Cardiomediastinum just refers to Enlarged Mediastinum, because Cardiomegaly is a separate label.

Annotation of simultaneous Cardiomegaly and Enlarged Cardiomediastinum will look like this:

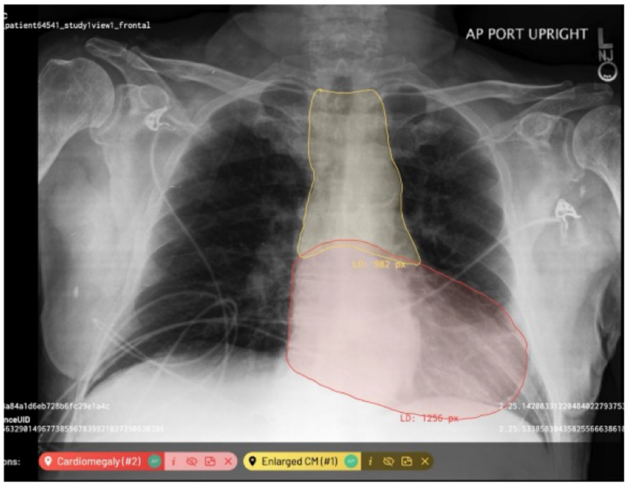

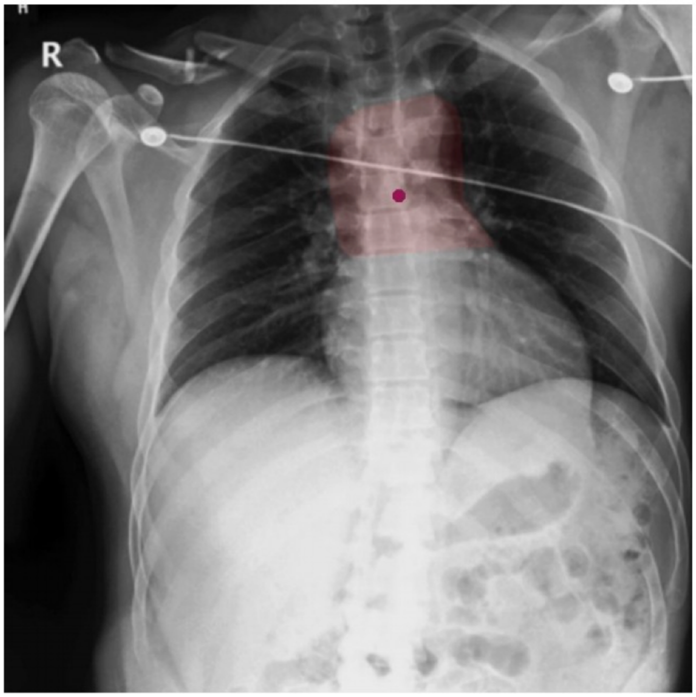

For Enlarged Cardiomediastinum, the most salient point will be in the center of the mediastinum.

**Supplementary Fig. S2 | Specific instructions given to benchmark radiologists on how to select the most representative point on a CXR for Enlarged Cardiomediastinum.**

#### Cardiomegaly

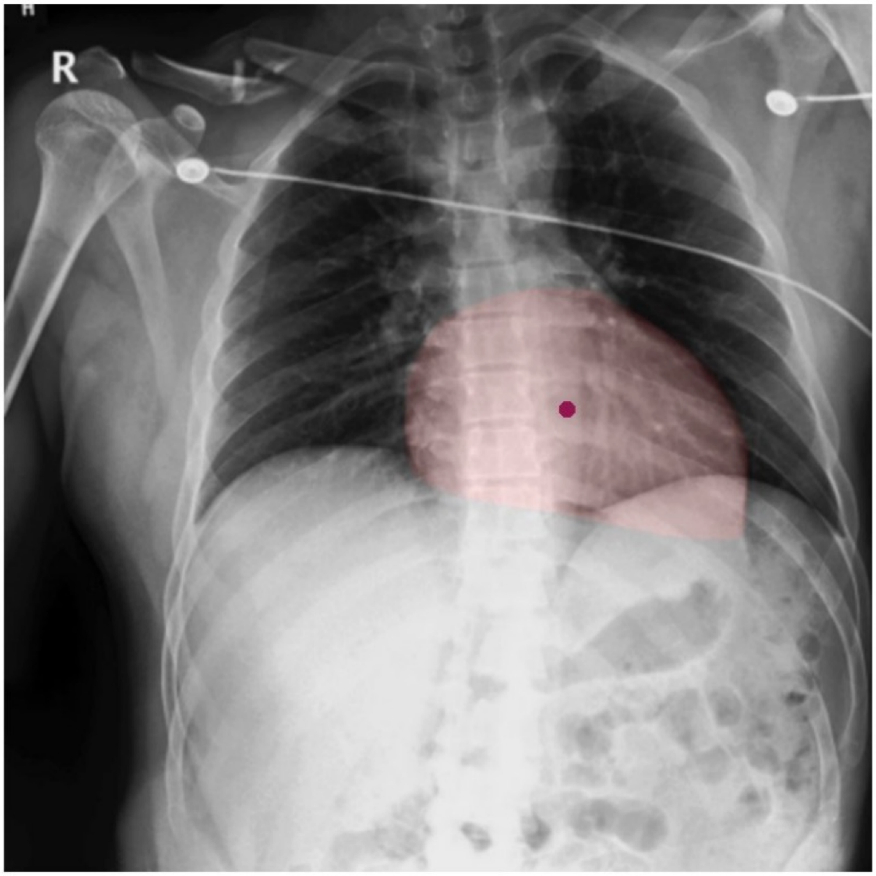

For Cardiomegaly, the dot should always be in the center of the segmentation of the enlarged heart.

**Supplementary Fig. S3 | Specific instructions given to benchmark radiologists on how to select the most representative point on a CXR for Cardiomegaly.**

#### Lung Opacity

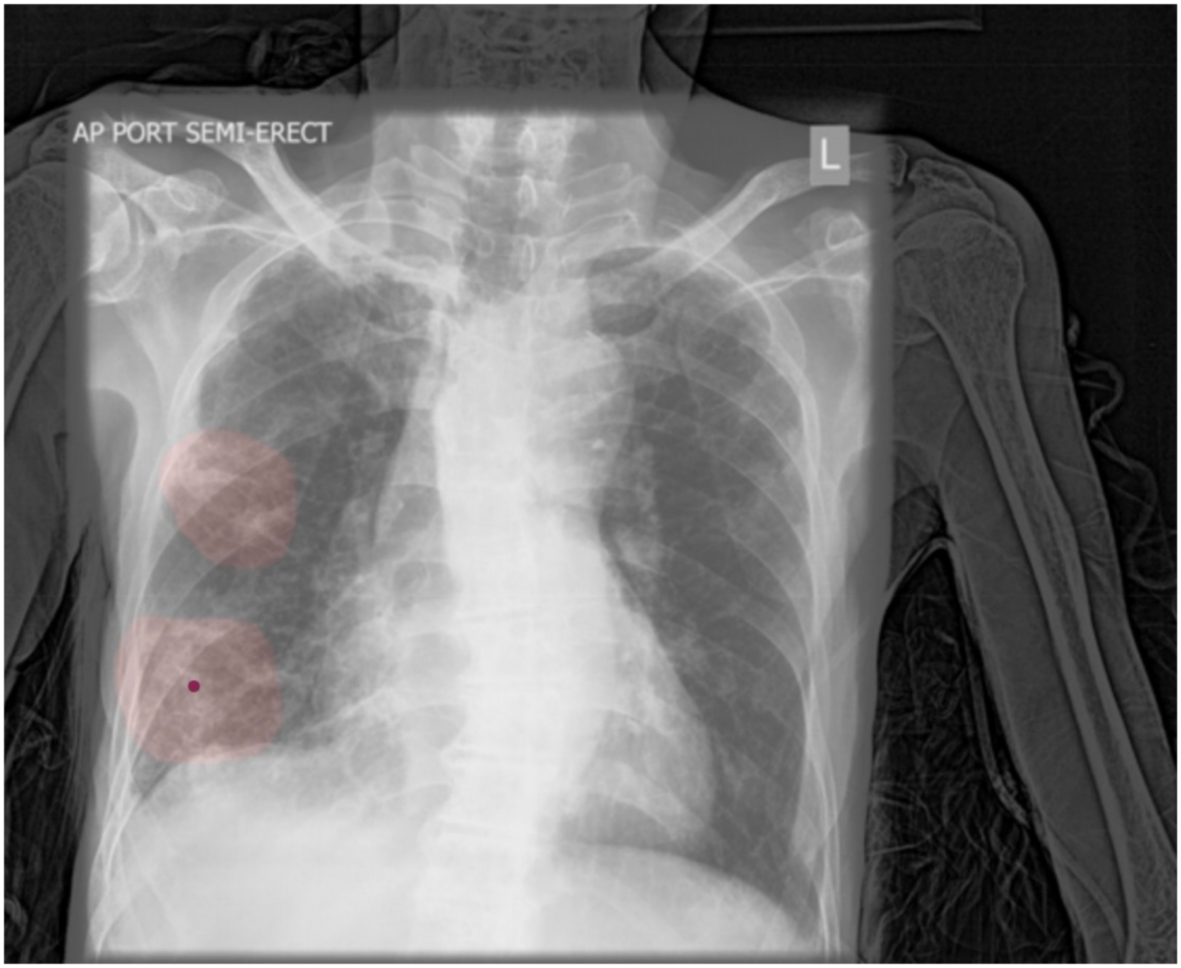

A Lung Opacity means a radiographic opacity that is not explained by the other types of opacities in our label set (Consolidation, Atelectasis and Lung Lesion). There may be more than one opacity. The most salient point should always lie in the center of the most prominent opacity.

**Supplementary Fig. S4 | Specific instructions given to benchmark radiologists on how to select the most representative point on a CXR for Lung Opacity.**

### Lung Lesion

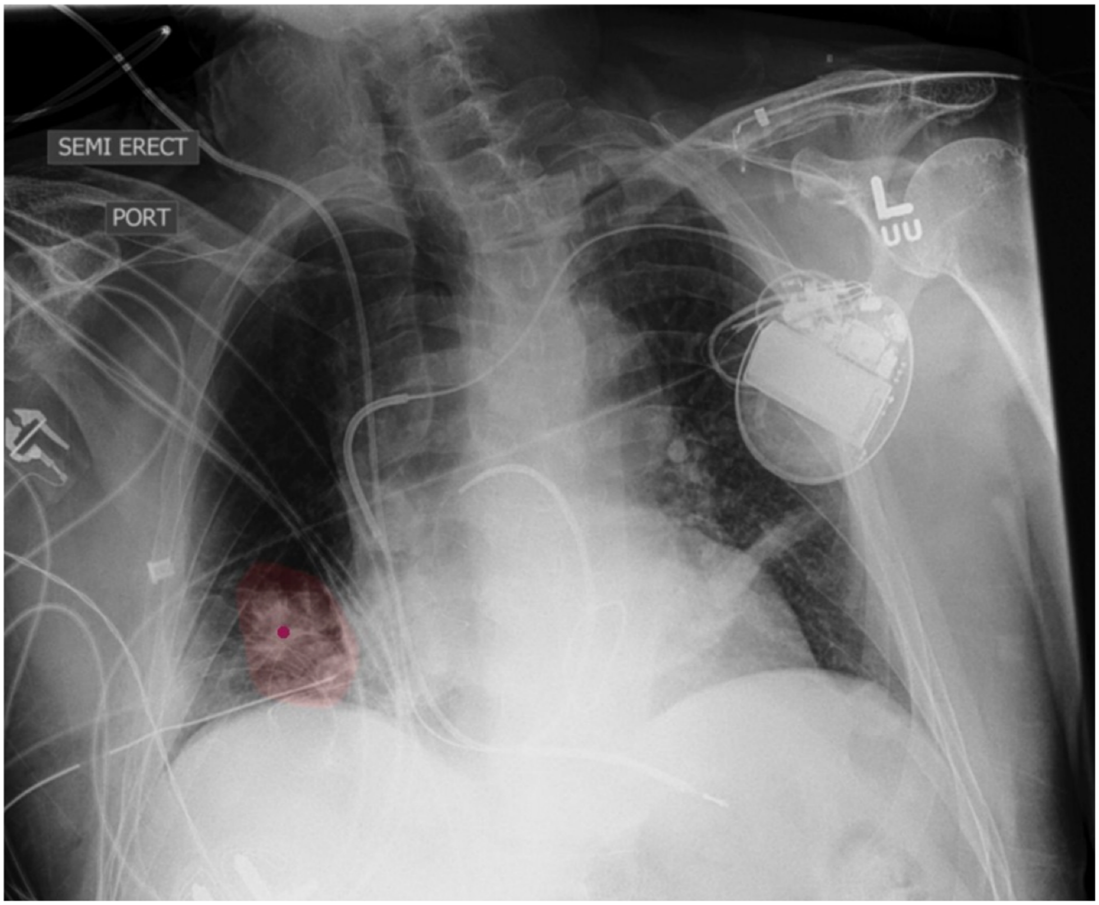

A Lung Lesion means a mass or nodule. There may be more than one lesion. The most salient point should lie inside the segmentation of the most pronounced lesion, and should be placed wherever that lesion is most pronounced.

**Supplementary Fig. S5 | Specific instructions given to benchmark radiologists on how to select the most representative point on a CXR for Lung Lesion.**

**Edema**

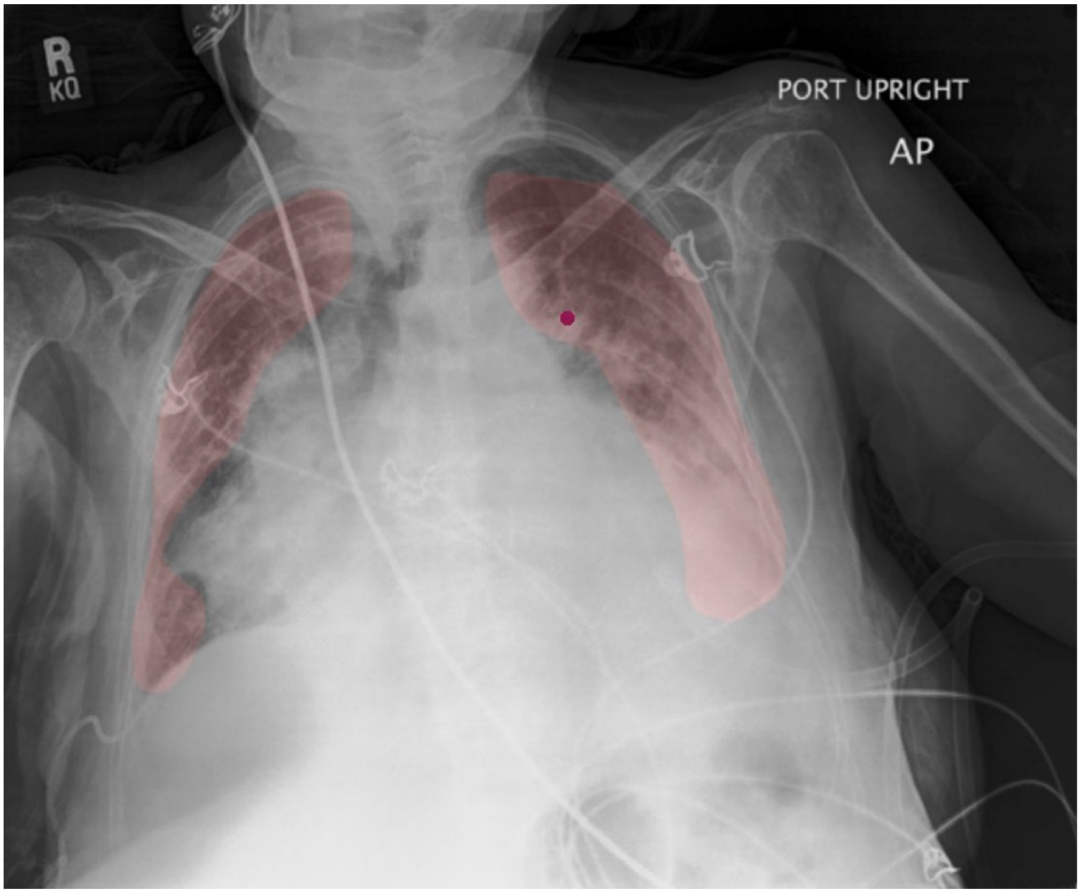

The most salient point should be placed wherever the visual features of edema are most pronounced. Please only select one side (right/left) where the visual features of edema are most pronounced.

**Supplementary Fig. S6 | Specific instructions given to benchmark radiologists on how to select the most representative point on a CXR for Edema.**

**Consolidation**

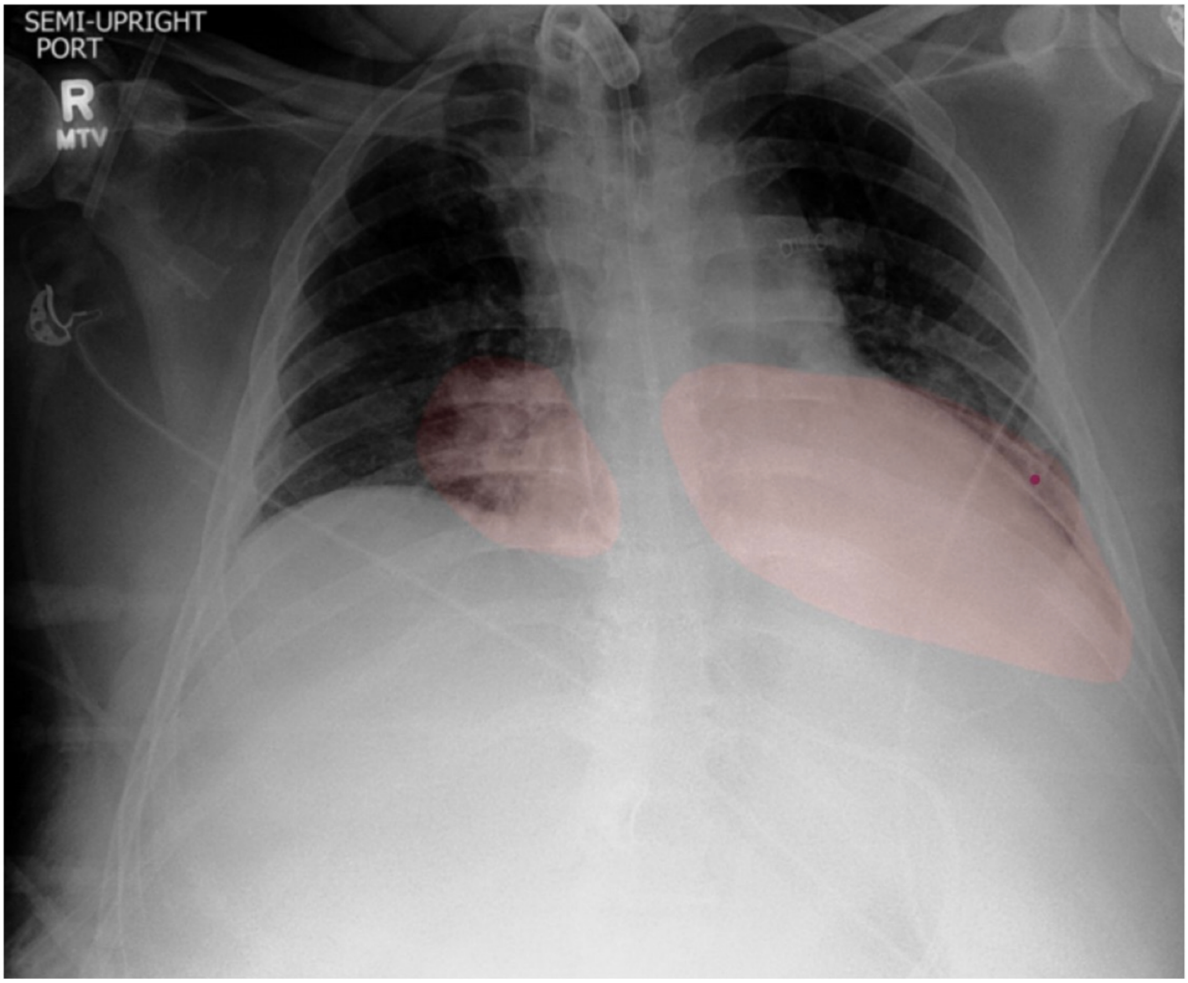

There may be more than one consolidation. The most salient point should lie inside the segmentation of the most pronounced consolidation, and should be placed wherever that consolidation is most pronounced. If the patient has diffuse consolidation throughout the entire lung field, aim for the area where the diffuse consolidation is more pronounced.

**Supplementary Fig. S7 | Specific instructions given to benchmark radiologists on how to select the most representative point on a CXR for Consolidation.**

### Atelectasis

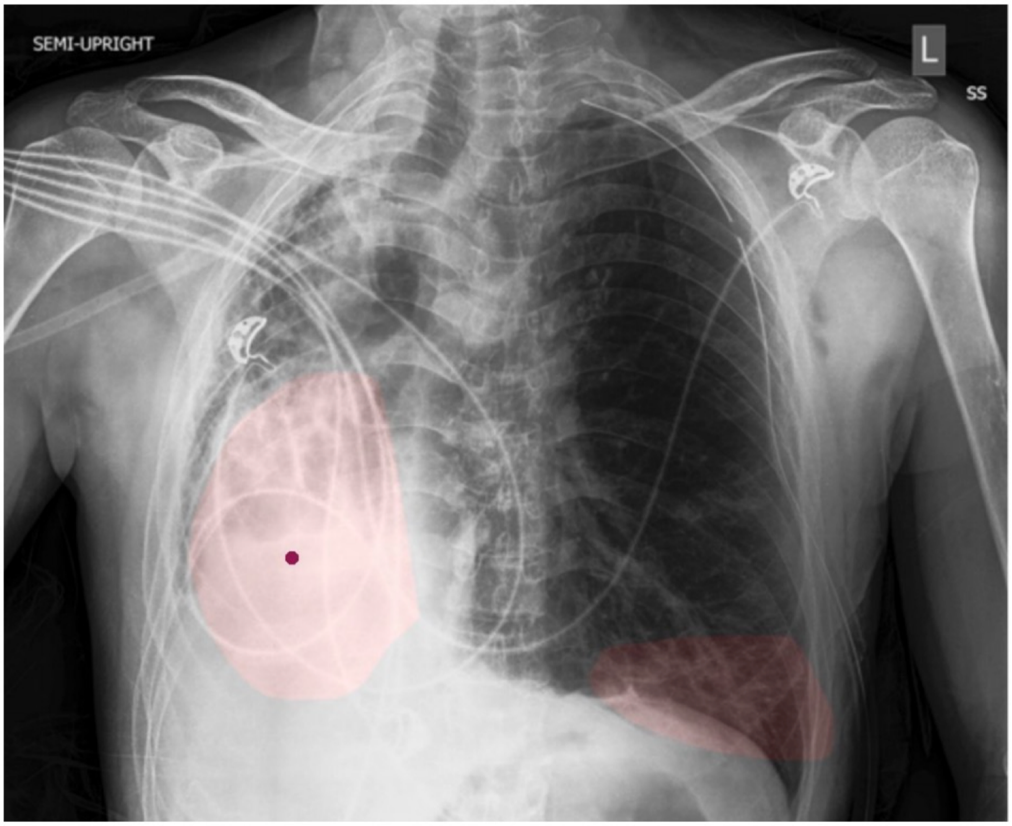

There may be multiple instances of Atelectasis. The most salient point should lie wherever the Atelectasis is most pronounced. As always, please only select one point, although there may be more than one segmentation.

**Supplementary Fig. S8 | Specific instructions given to benchmark radiologists on how to select the most representative point on a CXR for Atelectasis.**

**Pneumothorax**

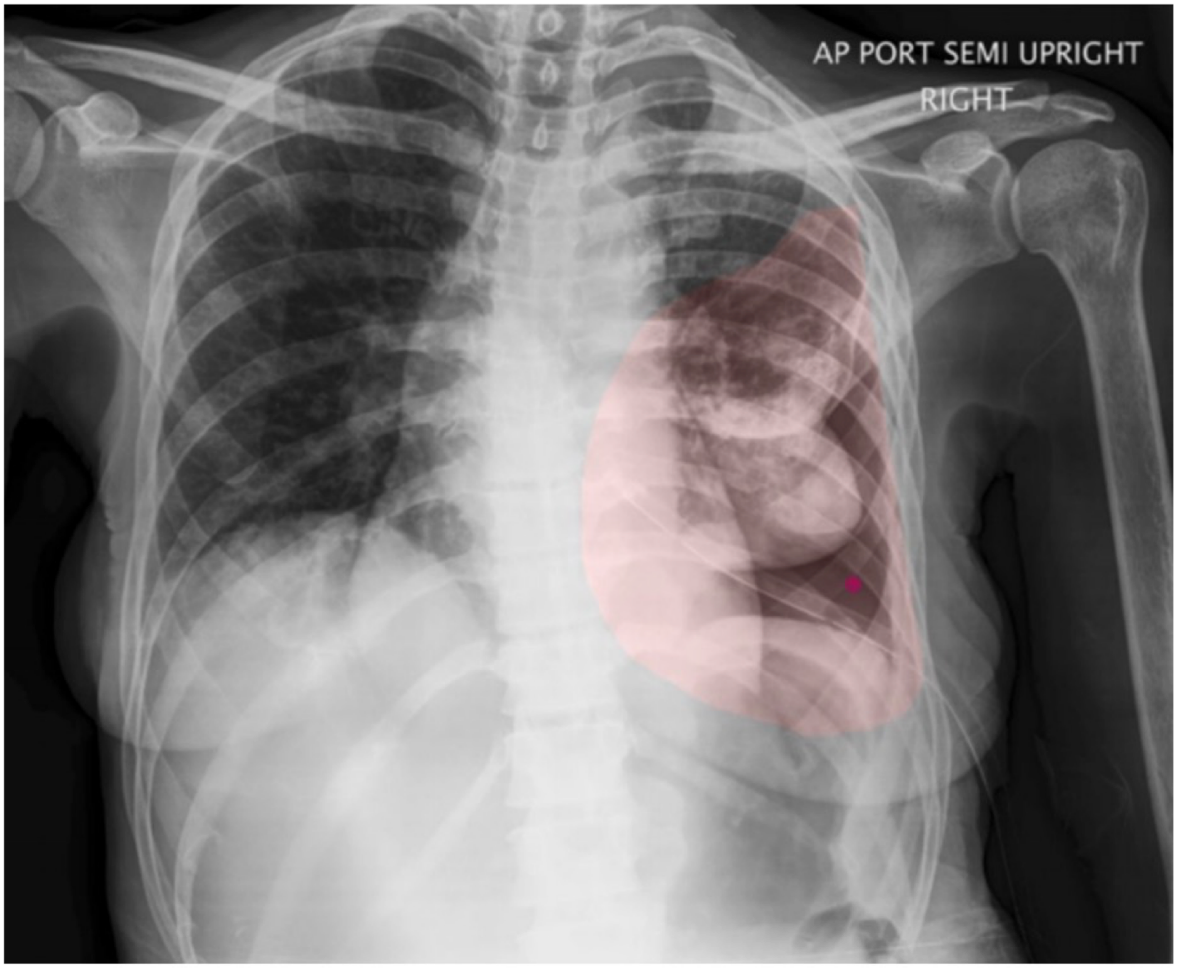

The single point should lie wherever the Pneumothorax is most pronounced.

**Supplementary Fig. S9 | Specific instructions given to benchmark radiologists on how to select the most representative point on a CXR for Pneumothorax.**

#### Pleural Effusion

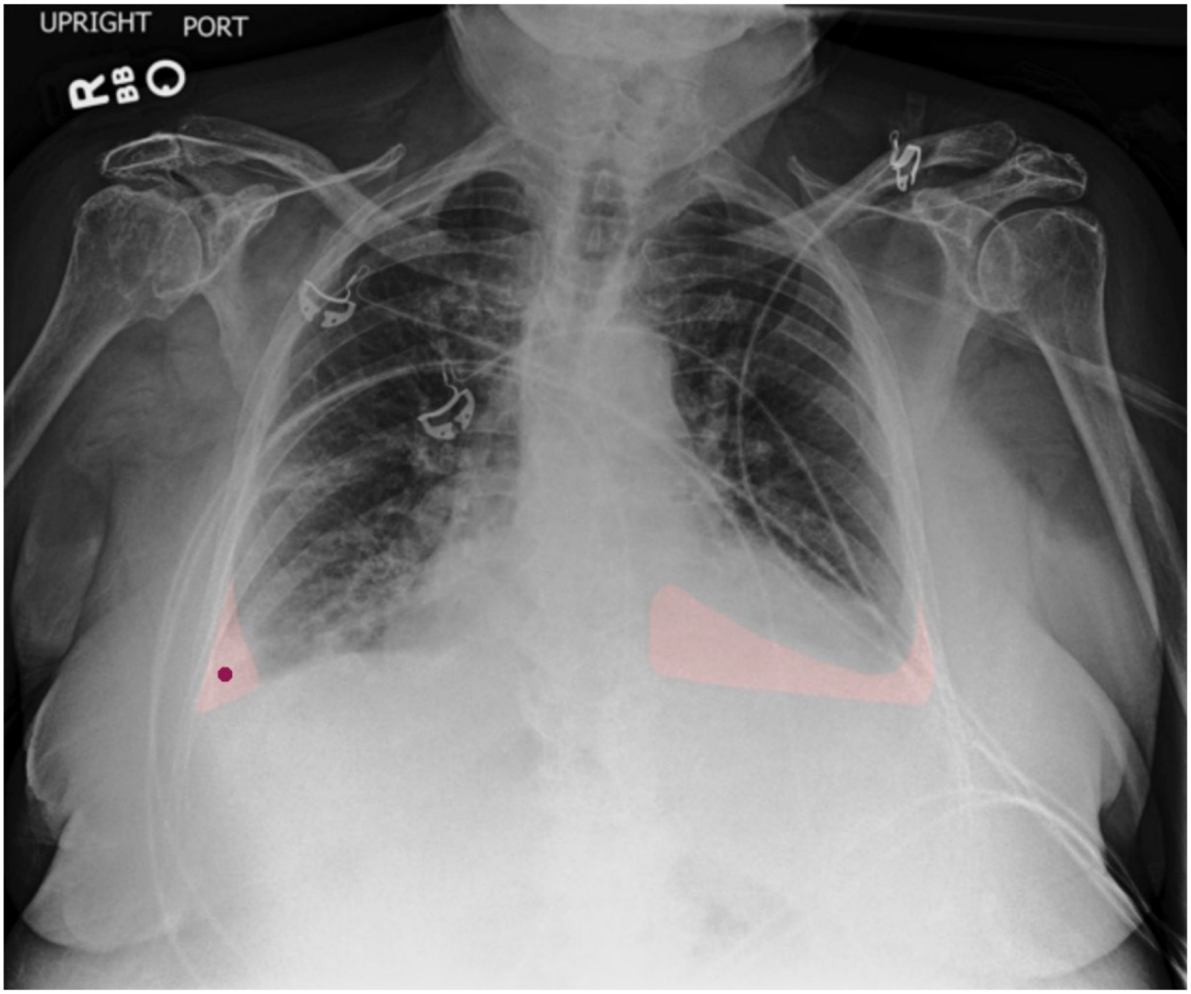

The most salient point should lie in the center of the pleural effusion. If the patient has bilateral pleural effusion, then please place the point in the center of whichever pleural effusion is more pronounced.

**Supplementary Fig. S10 | Specific instructions given to benchmark radiologists on how to select the most representative point on a CXR for Pleural Effusion.**

### Support Devices

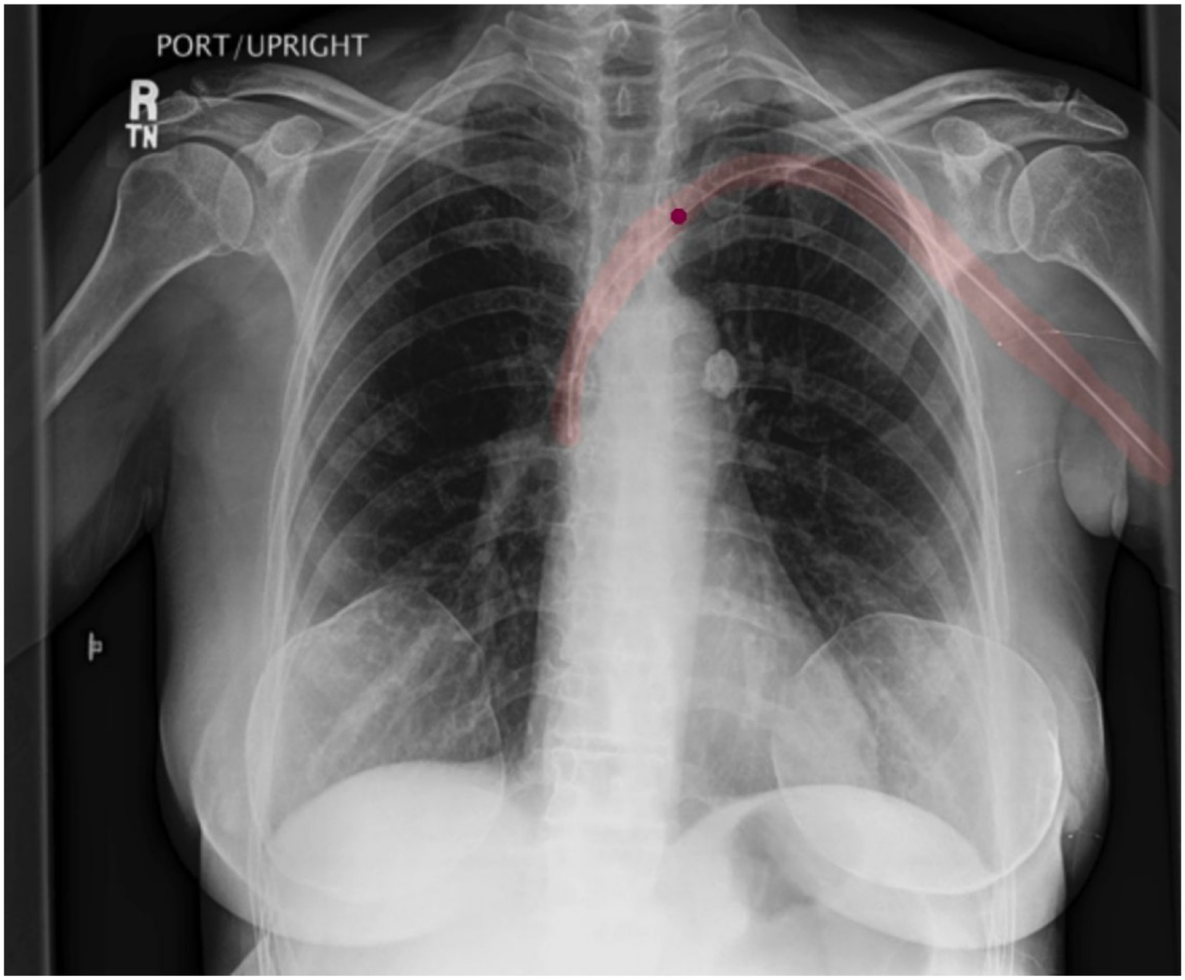

Support Devices are implanted or invasive devices such as pacemakers, PICC/central catheters, chest tubes, endotracheal tubes, feeding tubes and stents. ECG lead wires or stickers placed externally on the patient do not require labeling. If there is a single support device, please select the point at the estimated center of the support device. If there are several support devices, please select a point on the support device that you feel is most prominent. Please only consider the parts of the support device that are either inside (i.e. pacemaker, tube) or on (i.e. venous port) the patient. Do not place the point on a part of the support device that is completely outside the patient body (i.e. chest tube outside the thorax).

**Supplementary Fig. S11 | Specific instructions given to benchmark radiologists on how to select the most representative point on a CXR for Support Devices.**

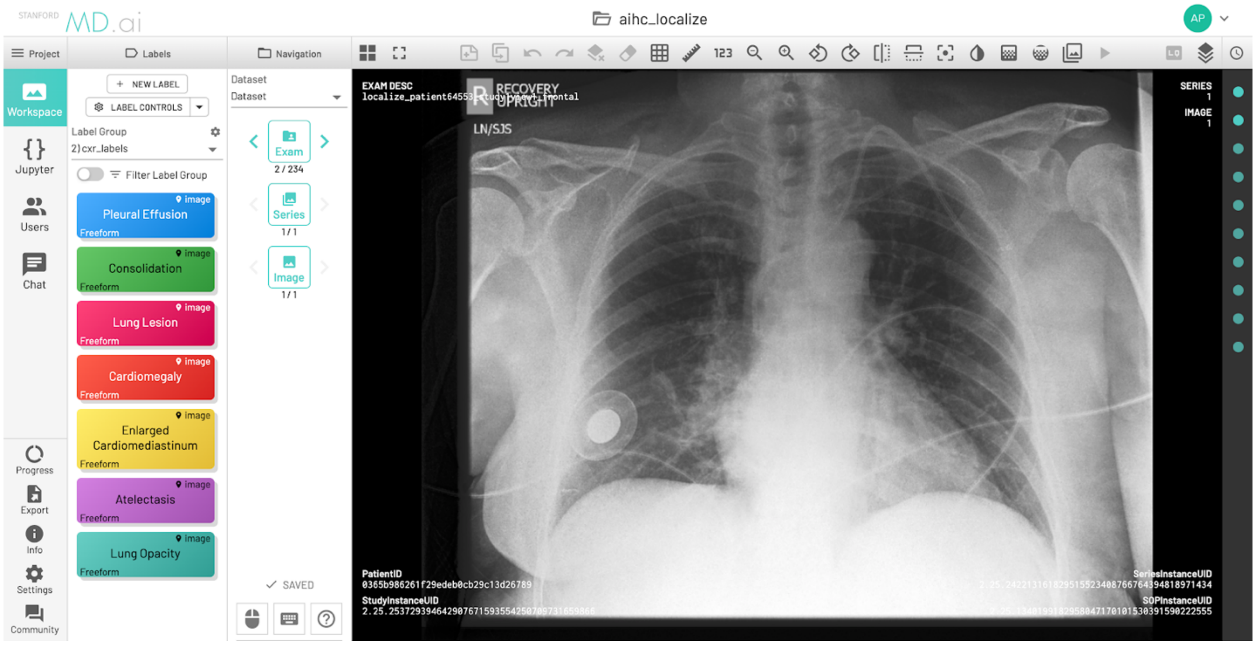

**Supplementary Fig. S12 | Screenshot of MD.ai project page where two radiologists drew ground-truth segmentations.** Radiologists could scroll through each CXR exam they were meant to segment. On the left side of the screen, a column of colored tabs, one for each of the 10 pathologies of interest.

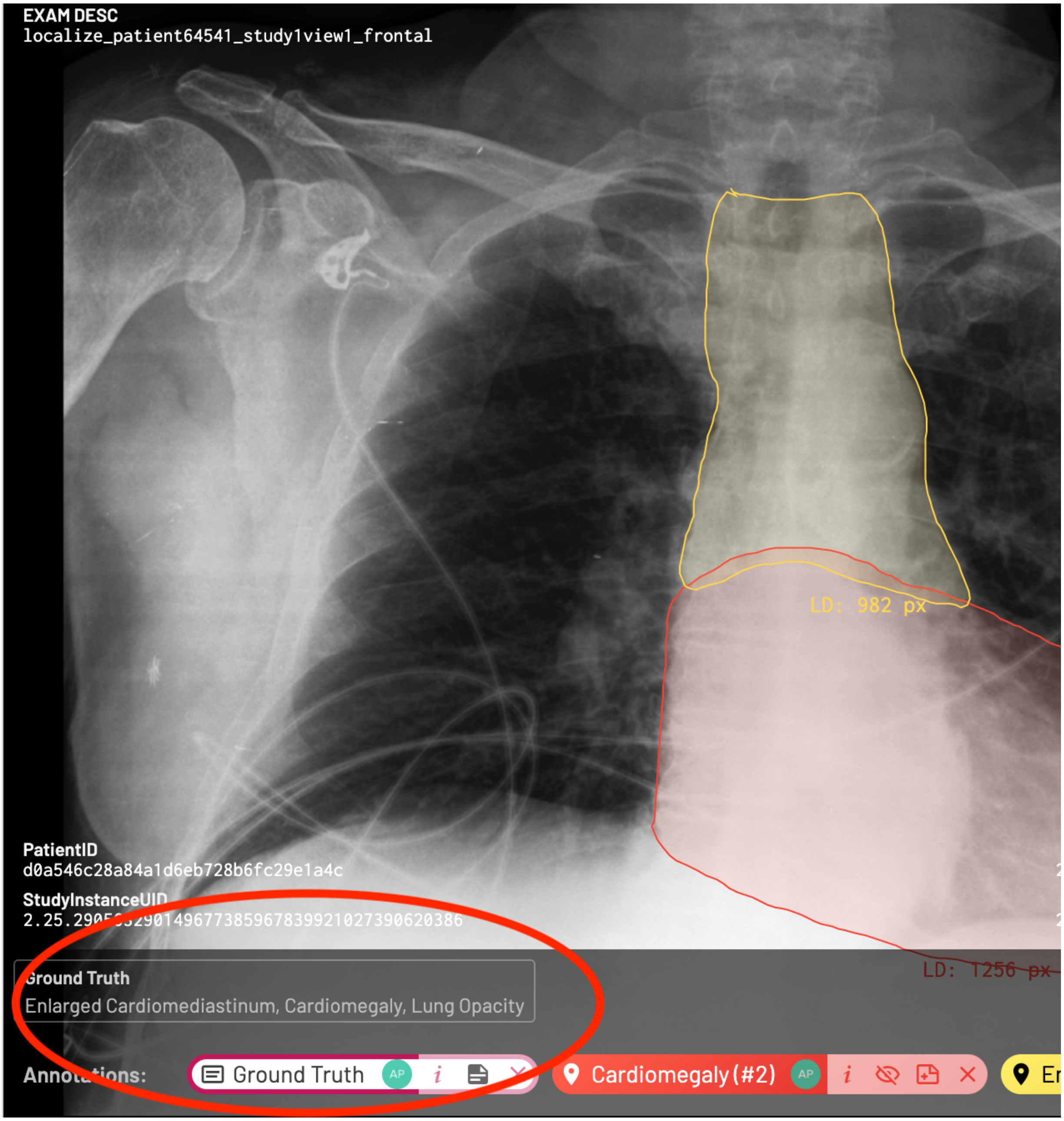

**Supplementary Fig. S13 | Screenshot of MD.ai ground-truth labels.** The radiologists had access to the ground-truth labels for each CXR, which were displayed at the bottom left of the CXR image.

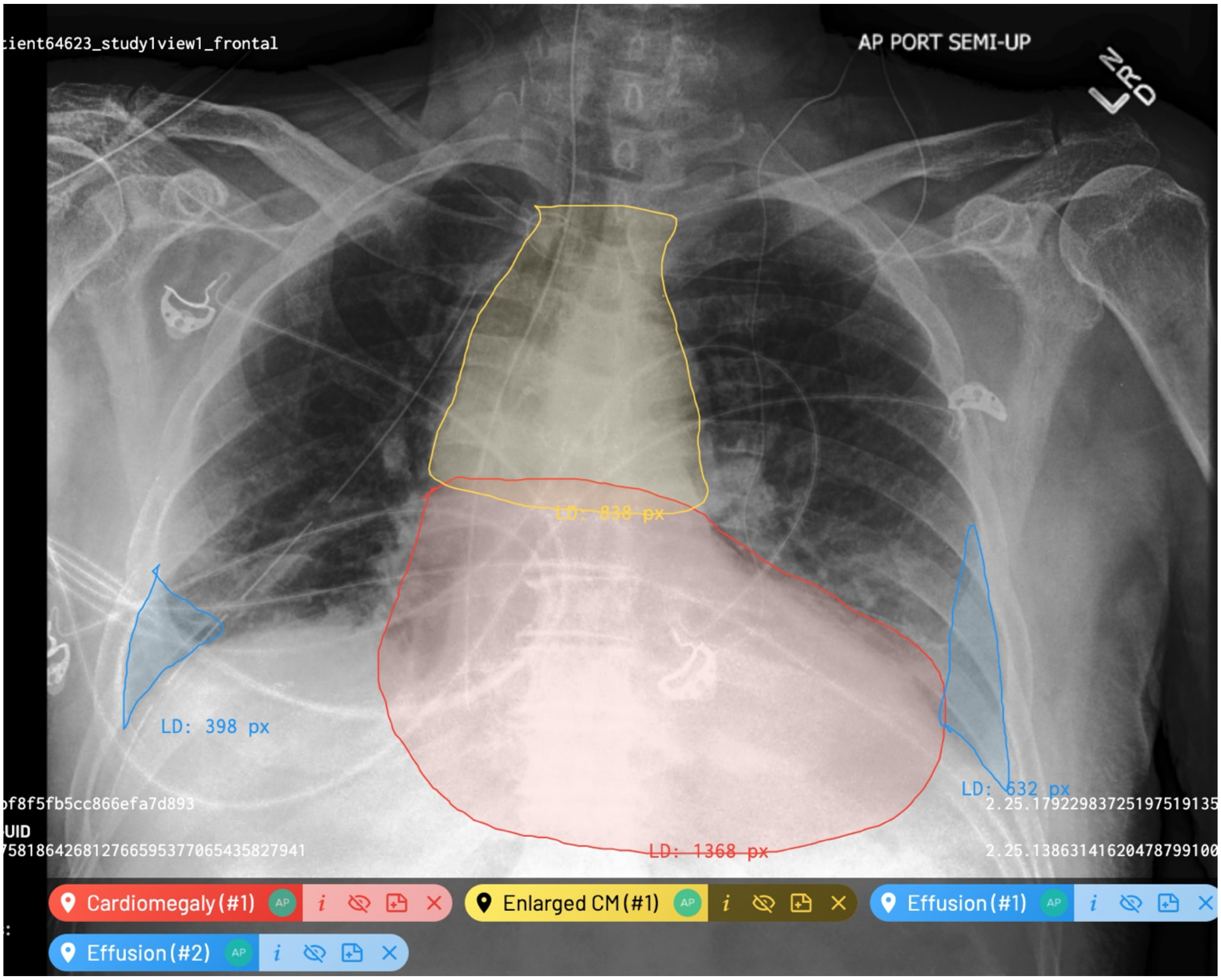

**Supplementary Fig. S14 | Screenshot of MD.ai segmentations for multiple pathologies on a single CXR.** Example CXR on MD.ai on which radiologists segmented Cardiomegaly, Enlarged Cardiomediastinum, and Pleural Effusion. After clicking on a pathology label from a sidebar on the left to activate the annotation mode, radiologists were able to draw directly on the CXR using free-hand contouring. After one pathology was segmented, the radiologist then segmented the next pathology in the same way. We asked the radiologists to strike a good balance between efficiency and accuracy.

**Supplementary Table S1 | Classification performance on test set.**

| Pathology | DenseNet121 | ResNet152 | Inception-v4 |
|---|---|---|---|
| Airspace Opacity | 0.926 | 0.923 | 0.912 |
| Atelectasis | 0.833 | 0.810 | 0.803 |
| Cardiomegaly | 0.885 | 0.879 | 0.863 |
| Consolidation | 0.868 | 0.862 | 0.876 |
| Edema | 0.915 | 0.901 | 0.897 |
| Enlarged Cardiom. | 0.583 | 0.609 | 0.578 |
| Lung Lesion | 0.912 | 0.900 | 0.869 |
| Pleural Effusion | 0.965 | 0.960 | 0.955 |
| Pneumothorax | 0.993 | 0.990 | 0.983 |
| Support Devices | 0.969 | 0.968 | 0.954 |
